# Data-driven risk/benefit estimator for multiple sclerosis therapies

**DOI:** 10.1101/2024.08.16.24312134

**Authors:** Bibiana Bielekova, Tianxia Wu, Peter Kosa, Michael Calcagni

## Abstract

**Background:** Multiple sclerosis (MS) disease-modifying treatments (DMTs) are tested in patients pre-selected for favorable risk/benefits ratios but prescribed broadly in clinical practice. We aimed to establish data-driven computations of individualized risk/benefit ratios to optimize MS care.

**Methods:** We derived determinants of DMTs efficacy on disability progression from re-analysis and integration of 61 randomized, blinded Phase 2b/3 clinical trials that studied 46,611 patients for 91,787 patient-years. From each arm we extracted 80 and computed 30 features to identify and adjust for biases, and to use in multiple regression models. DMTs mortality risks were estimated from age mortality tables modified by published hazard ratios.

**Findings:** Baseline characteristics of the recruited patients determine disability progression rates and DMTs efficacies with high effect sizes. DMTs efficacies increase with MS lesional activity (LA) measured by relapses or contrast-enhancing lesions and decrease with increasing age, disease duration and disability. Unexpectedly, as placebo arms’ relapse rate rapidly declines with trial duration, efficacy of MS DMTs likewise decreases quickly with treatment duration. Conversely, DMTs morbidity/mortality risks increase with age, advanced disability, and comorbidities. We integrated these results into an interactive personalized web based DMTs risk/benefit estimator.

**Interpretation:** Results predict that prescribing DMTs to patients traditionally excluded from MS clinical trials causes more harm than benefit. Treatment with high efficacy drugs at MS onset followed by de-escalation to DMTs that do not increase infectious risks would optimize risk/benefit. DMTs targeting mechanisms of progression independent of LA are greatly needed as current DMTs inhibit disability caused by LA only.

## Introduction

“Primum, non nocere!” (First, do not harm!)

This bioethics maxim reminds us that medical interventions have beneficial and harmful sides. Not only do risk/benefit ratios differ between subjects, in chronic diseases like MS they also change within subjects with the evolution of disease and comorbidities. Thus, we aimed to develop an accessible, user-friendly, and data-driven estimator of personalized risk/benefit ratios of MS disease-modifying treatments (DMTs) to enhance care for people with MS (pwMS).

We hypothesized that if sufficient cohort heterogeneity exists between clinical trials and we can correct systematic biases caused by decades of evolving MS diagnostics, treatments and trial designs, critical integration of trials data will identify patients’ characteristics that underlie both rates of MS disability progression and DMT efficacies. Resulting statistical models achieved high effect sizes and provide holistic view of MS and valuable insights on MS evolution and DMTs efficacy. Making therapeutic decision based on provided risk/benefits calculator could greatly improve MS outcomes.

## Methods

### Search strategy, trial inclusion criteria, PRISM diagram

The goal of the search strategy was to identify all published, randomized, blinded clinical trials that assessed efficacy of DMTs on the annualized relapse rate (ARR) and confirmed disability progression (CDP) in all forms of MS. The following criteria were used to screen search results:

1. The study contained at least 100 patients when adult MS patients were studied. We included pediatric MS trials if they had less than 100 patients because of the rarity of pediatric MS.
2. The study was blinded (double-blind or rater blinded).
3. The study was randomized.
4. The study reported Expanded Disability Status Scale (EDSS^1^)-based disability.
5. The study had minimum of 1 year/48 weeks treatment duration to be included in longitudinal analyses. Trials with shorter treatment duration were used for analysis of baseline parameters only.
6. To be included in longitudinal analysis of treatment effect, the study must have reported the proportion of patients with CDP based on EDSS change confirmed at a subsequent follow-up of 3 months/12 weeks or 6 months/24 weeks.

We started with 38 clinical trials identified in the 2017 meta-analysis of MS clinical trials^2^ (thus including the original search in the PRISM diagram; Supplementary Figure 1). We then added another PubMed search (on 3/2/2023) using the following inputs: Title/Abstract: “multiple sclerosis” AND “randomized” AND “clinical trial” AND “disability”. The search yielded 187 reviewed citations. We also searched clinicaltrials.gov database for keywords “multiple sclerosis” AND “randomized controlled trial”. The search yielded 347 studies of which 184 were completed. The clinicaltrials.gov identifier (NCT#) for the relevant studies was used to search PubMed (all fields). After excluding duplicates and studies not fulfilling inclusion criteria, we identified 61 unique trials testing 25 treatment modalities.

### Data extraction and imputation

For all treatment and control arms we extracted 80 data elements (Supplementary Statistical Workbook [SSW], Master tab; Supplementary Methods). We limited analyses to data elements included in a minimum of 10 trials because the power of statistical models depends on the number of trials with complete data. This excluded most volumetric MRI features. Differences in MRI methodology prohibited modeling remaining volumetric MRI features because we did not find relationships with moderate/high effect sizes (i.e., R^2^≥0.5) between the MRI volumetric data and cohort characteristics.

When supported by biological rationale and strong statistically significant relationships, we harmonized data elements. For example, 36 trials reported a baseline proportion of subjects with contrast-enhancing lesions (CELs%) on brain MRI and among these 25 (70%) also reported average numbers of CELs/scan (CEL#). Due to strong relationships between these parameters (R^2^ = 0.89, p<0.0001; Supplementary Figure 1), we used the resulting model to impute CEL% for trials that reported only CEL# and vice-versa. Analogously, while 79% of trials reported therapeutic efficacy on disability progression confirmed at 12 weeks (CDP@12wk), 13 trials also reported CDP confirmed at 24 weeks (CDP@24wk). We used the resulting linear regression model (CDP@12wk = [CDP@24wk + 1.5915]/0.9088; R^2^ = 0.98, p<0.0001; Supplementary Figure 2) to impute CDP@12wk for trials that reported only CDP@24wk.

During the modeling, we used models that explained ≥50% of variance to impute further missing values to maximize trial utilization. For example, for trials that did not report baseline ARR of the recruited population but did report baseline average CEL# we used *Eq#3* (R^2^=0.9; see results) to impute the baseline ARR. Likewise, some trials did not report disease duration (DD), which was selected as one of the efficacy predictors for *annualized* confirmed disability progression (A-CDP) in the multivariate model (i.e., *Eq#6*; results). Because we used *Eq#6* to compare efficacy between DMTs, we wanted to maximize the number of trials with DD. Therefore we construed a multivariate model to predict DD for relapse-onset MS (i.e., RRMS+SPMS, as trials missing DD recruited only relapse-onset MS). The DD model explained 84% of variance (p<0.0001 for all model predictors: DD = 2.9863*EDSS + 0.17043*%females – 12.8187; Supplementary Figure 3).

### Weighted stepwise multiple linear regression models

Due to modest number of trials, we used a 0.2 significance level to retain model confounders. We weighted trial results by the number of subjects (n) for cross-sectional and 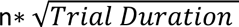 for longitudinal analyses^3^. To adjust for differences in trial duration, we recalculated EDSS^1^-based CDP rates to A-CDP, analogous to ARR.

The studentized residuals were used to examine the normality assumption of weighted stepwise multiple linear regression models and to verify the influential observations. We used the variance inflation factor (VIF < 4) to check multicollinearity.

### Analysis of the effect sizes of trials using active comparator

We observed that DMT efficacy depends on the recruited populations’ characteristics. Therefore, to estimate effect sizes against placebo in trials that used an active comparator, we predicted *average* DMT efficacy against placebo in the recruited population (using *Eq#6*). To estimate efficacy of a specific active comparator we computed its weighted *average* residual from observed versus *Eq#6*-predicted efficacies using its placebo-controlled trials. Finally, measured progression rates and predicted comparator efficacy (against the recruited population’s placebo) predicted progression rates in the “in-silico placebo arms” of active comparator trials.

**Penalization function for trials that biased inclusion criteria against active comparator and analyses of dynamic efficacy data from digitalized Kaplan Meier (K-M) curves** are described in the Supplementary Methods and Results.

### Data availability

All extracted/computed data are in the SSW Master sheet, sheets s1-11 contains SAS exports for results, and sheets s12-14 contain mortality tables.

## Results

The paper has two parts: 1. Estimating efficacy of DMTs on MS disability progression based on a patient’s baseline characteristics; 2. Estimating the patient-specific risk of morbidity/mortality from infections and cancers due to MS DMTs.

Because we did not identify suitable real-world data for estimating personalized efficacy of MS DMTs, we used randomized, blinded, and controlled clinical trials.

### Study population

We analyzed 131 arms from 61 randomized, blinded, controlled clinical trials studying 46,611 MS patients for 91,787 patient-years. For untreated MS analyses, 9,165 patients in placebo arms were studied for 19,410 patient-years.

### Clinical trial inclusion/exclusion (I/E) criteria enrich for subjects with a favorable risk/benefit ratios

All 42 relapsing-remitting MS (RRMS) trials were enriched with subjects with LA by requiring evidence of relapse or CEL in the past two years. 98% of RRMS trials excluded subjects requiring a walking cane (i.e., EDSS > 5.5) and 95% imposed age limits, with 83% excluding subjects older than 55 years.

84% of Progressive MS (PMS) trials excluded subjects older than 65 years. All PMS trials restricted maximum disability: 79% excluded non-ambulatory subjects (EDSS>6.5).

All trials excluded subjects with cancers and infections.

Restricting disability levels and requiring LA evidence decreased the recruited population’s mean age. I/E criteria predicted the recruited population’s age (R^2^=0.83; Figure 1A), even when omitting maximum age restriction (R^2^ = 0.80; Figure 1B, SSW-s1).

**Figure 1:**
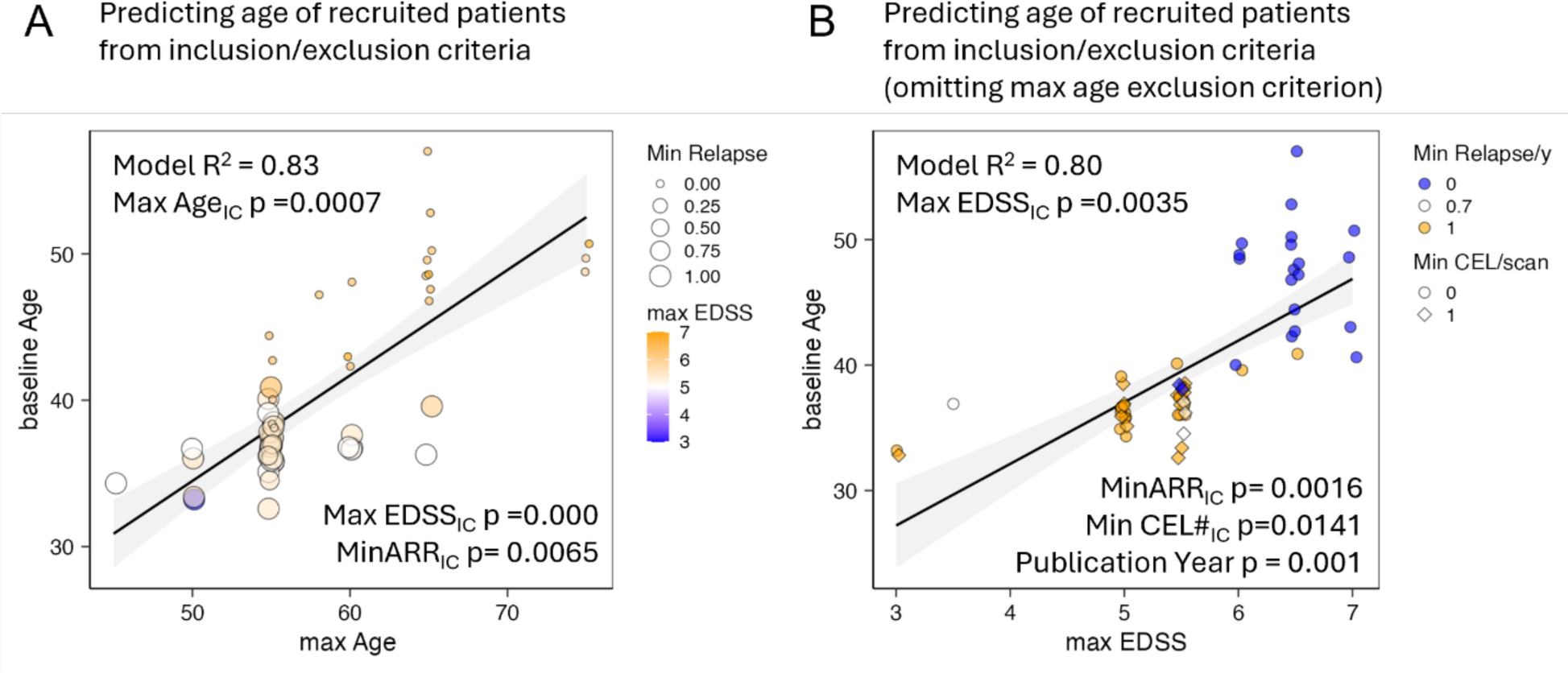
Inclusion/exclusion (I/E) criteria of MS trials reliably predict age of the recruited population even if ignoring upper age limit restriction. **A.** I/E criteria of maximum age, maximum EDSS and Minimum ARR (number of relapses in preceding 1-2 years) predict baseline age of the recruited population with R^2^=0.83 in stepwise multiple regression model. We displayed I/E of maximum age on x-axis, maximum EDSS as color heatmap (small EDSS in blue, high in orange) and minimum ARR as size of the circles. The R^2^ of the model and p-values of the predictors are displayed in the plot. **B.** Even when omitting I/E of maximum age altogether, the remaining I/E criteria effectively determine age of the recruited population. I/E criterium of maximum EDSS is on the x-axis, min ARR is shown by color scheme (Minimum ARR 1 in orange, minimum ARR 0 in blue) and minimum CEL# as shape, with minimum CEL# ≥1 as rhomboids and CEL#=0 as circles. The R^2^ of the model and p-values of the predictors are displayed in the plot.

We conclude that trials are enriched with subjects with favorable risk/benefit ratios. To assume that results apply to patients at the criteria’s maximum age is misleading because, due to remaining I/E criteria, older subjects represent a negligible (undisclosed) proportion of the studied population.

### Newer trials recruit patients with more benign MS

Trial recruitment dates may affect outcomes due to changes in trial design, MS diagnostic criteria and DMT availability.

With increasing publication year used as a proxy for recruitment dates, untreated MS patients (i.e., placebo cohorts) exhibited decreasing ARR (R^2^=0.6 for relapse-onset MS: i.e., RRMS + SPMS and R^2^=0.76 for RRMS only, p<0.0001 for both; Figure 2A, SSW-s2). Likewise, A-CDP rates decreased in untreated patients with increasing publication year (R^2^=0.46, p=0.0003 for relapse-onset MS and R^2^ = 0.36, p=0.0143 for RRMS only; Figure 2B, SSW-s3).

**Figure 2:**
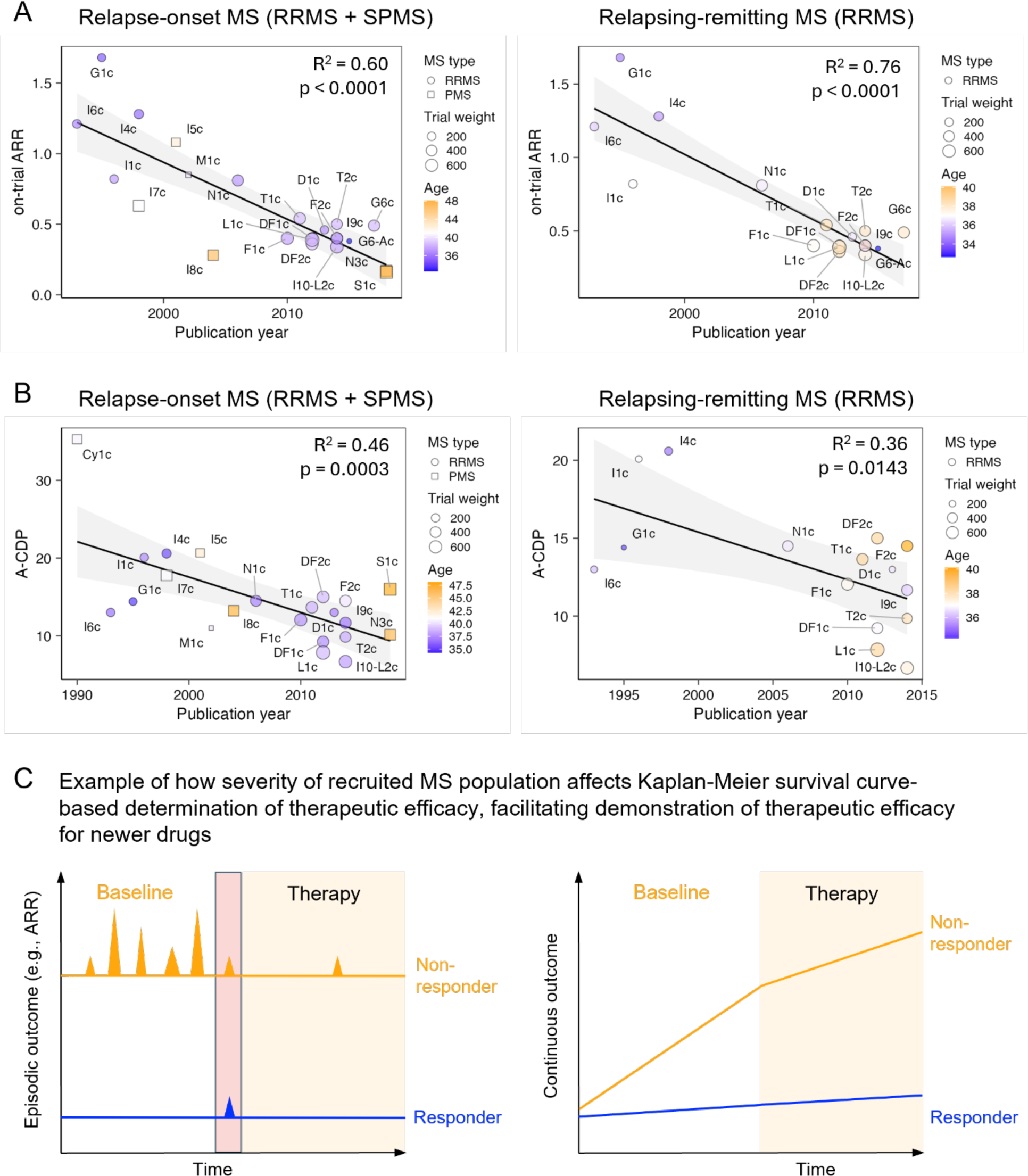
Newer trials recruit patients with more benign MS, which likely advantages newer over older drugs in comparative efficacy. **A.** Relationship between on-trial measured ARR and publication year in placebo arms of relapse-onset (i.e., RRMS+SPMS) trials (left panel) or RRMS only trials (right panel). The R^2^ and p-values for the effect of Publication year are displayed in panels. RRMS trials are shown as circles and progressive MS trials as squares (with size proportional to trial weight). Age is represented with heatmap color. **B.** Relationship between on-trial measured annualized EDSS-based confirmed disability progression (A-CDP) and the publication year in placebo arms of relapse-onset trials (left panel) or RRMS only trials (right panel). The R^2^ and p-values for the effect of Publication year are displayed in panels. RRMS trials are shown as circles, progressive MS trials as squares, with size proportional to trial weight. Age is represented with heatmap color. The trial name corresponds to the Index column in the Supplementary Master Worksheet (SSW). **C.** Conceptual model that explains how recruiting patients with more benign MS may provide advantage to newer over older drugs in comparative efficacy: MS trials dichotomize outcomes (i.e., patient did or did not experience relapse, patient did or did not progress), ignoring the severity of the relapse or severity of progression. It is easier for a DMT to completely suppress episodic outcomes (e.g., relapse activity or EDSS progression) or continuous outcomes (e.g., increase in T2 lesion load) during short trial duration for patients with benign MS (depicted in blue) in comparison to patients with aggressive MS (depicted in orange).

Figure 2 shows unexpectedly high effect size that publication year exerts: compared to 1990s trial populations, recently recruited patients have approximately 60% fewer relapses and 40% lower A-CDP rates. This bias likely impacts reported efficacy, because MS trials measure efficacy by dichotomizing patients into those who did or did not progress and patients with severe disease more likely experience breakthrough disease activity and progression on treatments (Figure 2C). We adjusted for this bias in downstream analyses.

### MS LA strongly decreases with age

MS LA is measured by the CEL# and ARR. In the baseline cohort characteristics, we observed significant decrease of LA with age, predicting 0 CEL# at mean age of 59 years (Figure 3A, SSW-s4) and 0% ARR after age of 50 years (Figure 3B, SSW-s5).

**Figure 3:**
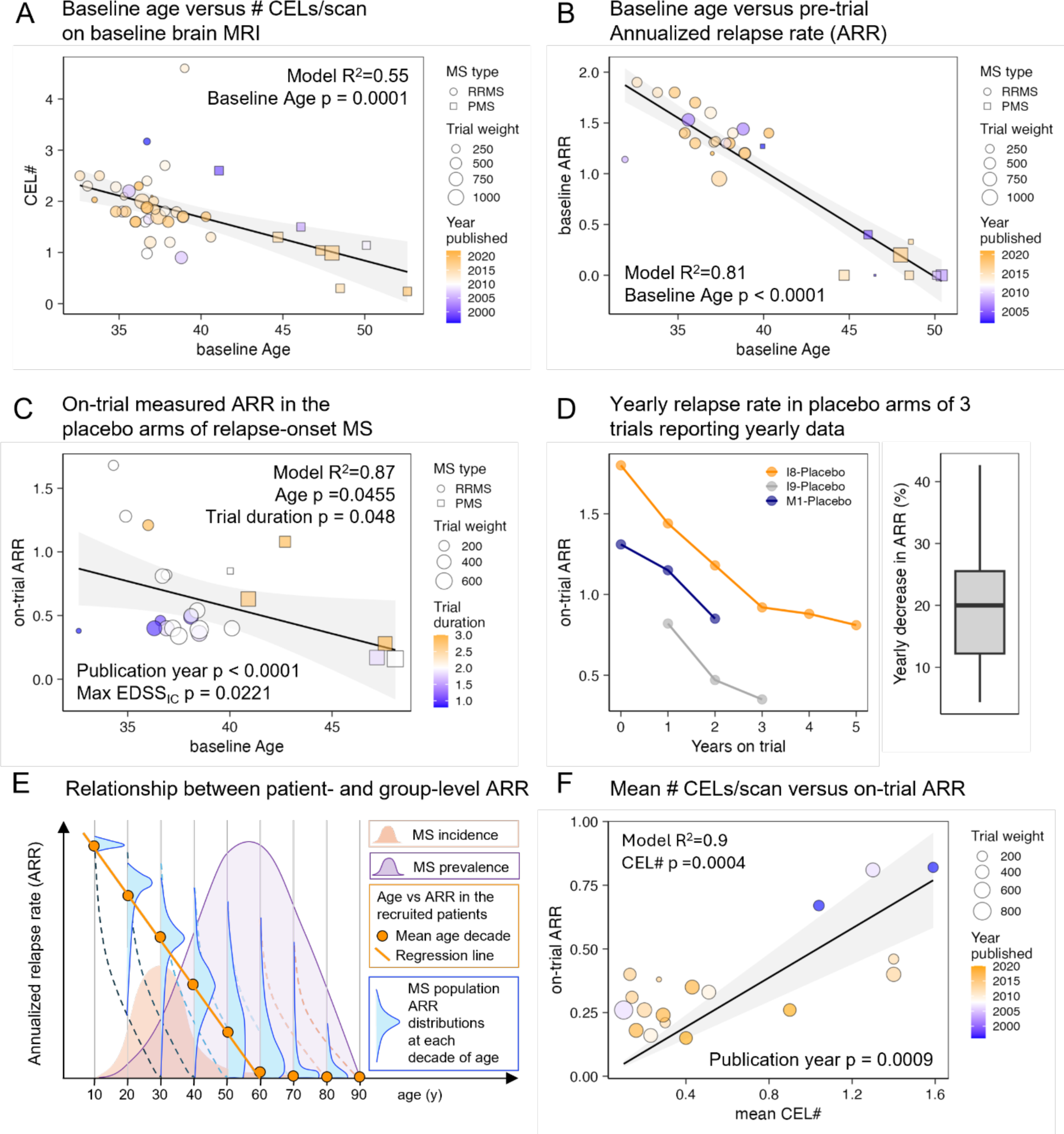
MS LA decreases with time measured as age and trial duration. **A.** Mean CEL# per baseline brain MRI scan decreases with the mean baseline age of the recruited population in relapse-onset MS (i.e., RRMS + SPMS). Only 1 arm/trial was used for each trial that reported both variables and the weight is the number of patients in this arm. R^2^ of the stepwise multiple regression model and p-value of Age predictor are displayed. RRMS trials are displayed as circles and SPMS trials as squares, with size proportional to weights. Publication year is displayed as color heatmap with older trials in blue and newer trials in orange. **B.** Baseline Annualized relapse rate (ARR) decreases significantly with the mean baseline age of the recruited population (ARR zero was imputed for trials that recruited only primary progressive MS (PPMS), as by definition these patients do not experience relapses. Only 1 (larger) arm/trial was used. R^2^ of the model and p-value of Age predictor are displayed. **C**. Because panels A and B are biased by inclusion/exclusion criteria requiring LA for relapse-onset MS, we studied on-trial measured ARR in placebo arms of relapse onset MS. Trial weight is number of subjects in placebo arm 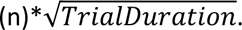 Trial duration is also displayed as color heatmap with shorter trials in blue and longer trials in orange. R^2^ of the model and p-value of Age predictor are displayed. **D.** Three trials (trial index I8, I9 and M1, Index column in the SSW Master Worksheet) reported ARR for placebo arms for each trial year, showing on-trial yearly decrease by about 20%. Right panel shows box blot of all available yearly changes from these trials; box displays 1^st^ and 3^rd^ quartiles, horizontal line is median and whiskers are minimum and maximum). Year 0 are baseline data for recruited patients that were not reported for I9 trial. **E.** Conceptual model that reconciles apparent slower decrease in ARR with age as compared to faster decrease with trial duration. The on-trial measured ARR decline in the placebo arms displayed in panels C and D represent average within-subject decrease in ARR and reflects the true rate of ARR decline in untreated MS. The seemingly slower population level decline measured by baseline age is biased: inclusion criteria preselected from each age category only those patients who have relatively short disease duration and therefore high LA reflected by ARR and CEL#. The smaller orange histogram shows MS incidence peaking around age 30 years with right tail of rare patients with late MS onset. Small vertical blue histograms represents distribution of ARR for all MS patients of that age (displayed for each age decade), with age-population median shown as orange dot. The rapid decline in ARR for patients of the same (decade) age is shown as dashed lines while population level decline (affected by constant contribution of newly diagnosed patients recruited to trails because of their LA) as orange solid line. Purple MS prevalence histogram is adopted from USA MS prevalence data reported by Wallin et al^27^ and shows that most patients living with MS are excluded from MS clinical trials by inclusion/exclusion criteria. Additionally, even when using biased estimated of MS LA in relationship with age displayed in panels A and B, it is clear that most patients living with MS lack MS LA. **F.** Because ARR is no longer measurable in contemporary clinical practice when MS DMTs should be initiated after first (correctly diagnosed) MS attack we used on-trial measurements of mean CEL# and on-trial ARR from all trials/arms that reported these data to investigate if CEL# can predict ARR. We forced model intercept to zero, as negative numbers of ARR or CEL# cannot exist. Trial weight is 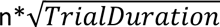 and publication year (significant covariate in the model) is displayed as color heatmap with older trials in blue and newer in orange colors. Model R^2^ and p-value of CEL# predictor are displayed.

These estimates are biased by LA inclusion criteria. Placebo arms measure true declines of LA. On-trial ARR decreased with increasing age of relapse-onset MS ([*Eq#1*], Figure 3C; R^2^=0.75, p<0.0001; SSW-s6). Publication year also decreases ARR (p<0.0001):

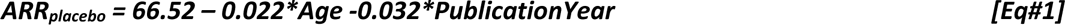

Baseline age likewise negatively correlates with ARR in the RRMS only model (R^2^=0.87, p<0.0001; [*Eq#2*], SSW-s6):

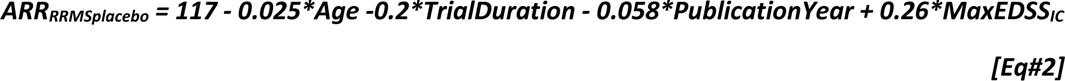

Eq#2 indicates that ARR in untreated RRMS patients decreases with age (p=0.046) and trial duration (p=0.048). This indicates rapid ARR decline, measurable within (short) trial duration. Three trials reported yearly placebo arms relapse rates (Figure 3D). The within-trial data confirm unexpectedly rapid (∼20% yearly) ARR decline, which is consistent with *Eq#2* prediction. Figure 3E reconciles an apparent discrepancy between rapid intra-individual declines (placebo arms) with slower population ARR declines.

Unfortunately, ARR is unobtainable for most patients in contemporary practice because they are (ideally) diagnosed and treated soon after first relapse. As the MS diagnostic process includes contrast-enhanced MRI, we observed that CEL# predicts ARR (Figure 3F; [*Eq#3*], R^2^=0.9, CEL# p=0.0004, Publication Year p=0.0009; *SSW-s7*) and can be used as an ARR surrogate.

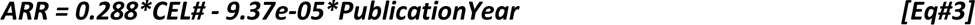

### Annualized EDSS-based confirmed disability progression rate (A-CDP) in untreated MS increases with ARR and disease duration (DD)

Stepwise regression using age, DD, EDSS, proportion of females in trial population, measured ARR, trial duration and publication year identified predictors of placebo arms A-CDP [*Eq#4*]. The model explains 57% of variance and includes only ARR (p=0.0014) and DD (p=0.0072; Figure 4A; SSW-s8).

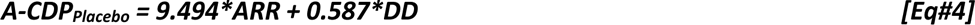

**Figure 4:**
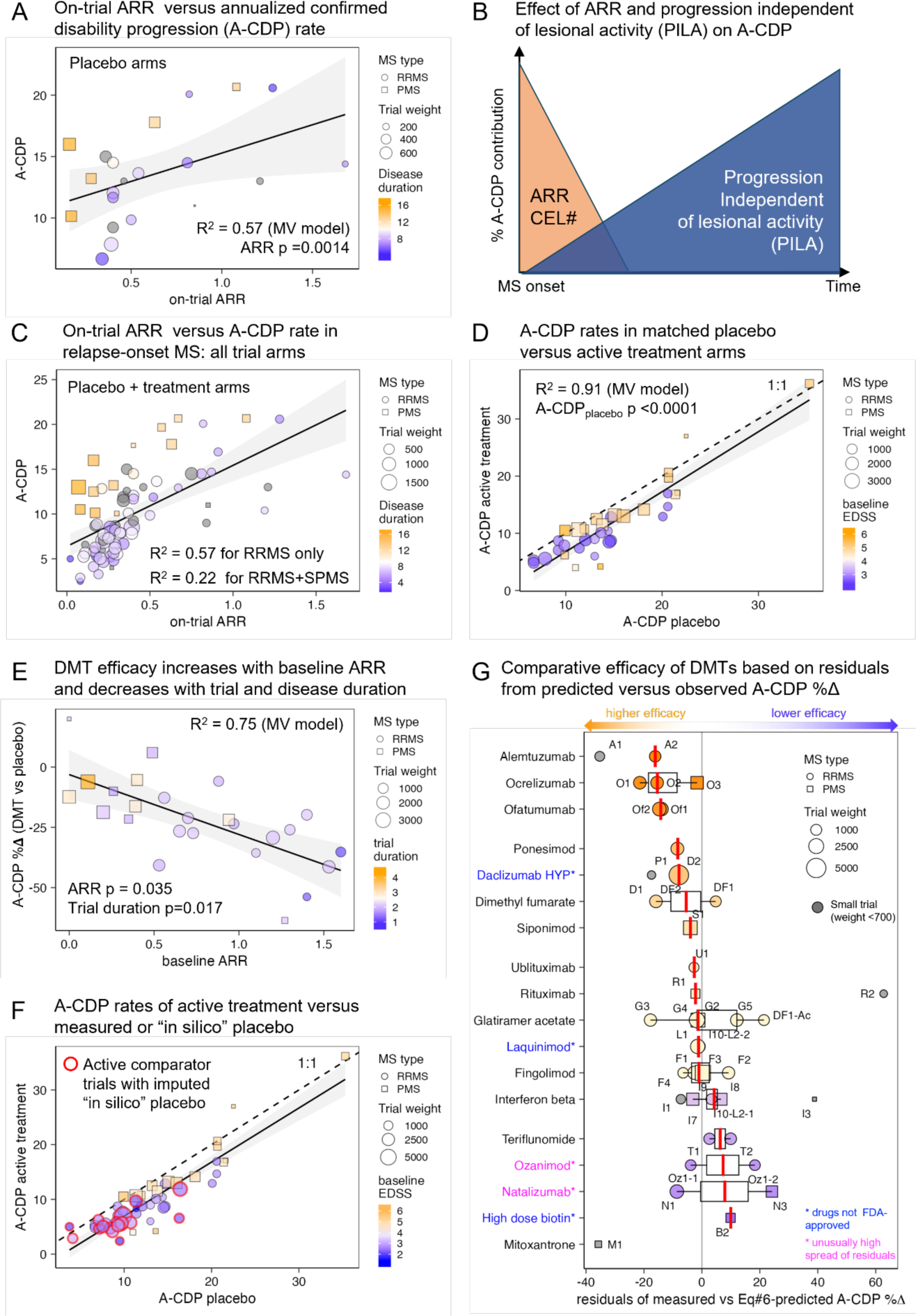
Baseline characteristics of MS population (i.e., ARR, CEL#, EDSS, DD) are stronger predictors of the annualized confirmed disability progression (A-CDP) than treatment status, and DMTs efficacy increases with ARR and decreases with treatment duration. **A.** On-trial measured ARR versus A-CDP in the placebo arms of relapse-onset MS. For all panels, RRMS trials are displayed as circles and SPMS trials as squares, with size proportional to trial weight 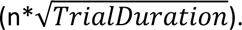 Mean DD is coded according to color heatmap (shorter in blue, longer in orange). The model R^2^ and p-values of model predictors are displayed. **B.** Conceptual representation of the two opposite contributors to A-CDP: MS LA displayed in orange color that is highest at MS onset and rapidly decreases thereafter and Progression Independent of LA (PILA) displayed in blue color that increases steadily with DD. **C**. On-trial measured ARR versus A-CDP for all arms of relapse-onset MS, analogous to panel A. The significant correlation between ARR and A-CDP is clearly observed for both RRMS and PMS trials/arms with R^2^ of this relationship displayed separately for RRMS trials and all trials. DD codified by color heatmap (shorter blue, longer orange) explains residual variance. **D.** Relationship between A-CDP reported in the placebo and active treatment arms. Dotted line represents 1:1 line, showcasing that on average, active treatment arms accumulate disability at slower pace than placebo arms, but the difference is small. The major determinant of progression rates are baseline characteristics of the recruited population. Baseline EDSS is coded according to color heatmap (low disability blue, high disability orange), showing that populations with higher baseline disability experience lower therapeutic efficacy. The model R^2^ and p-value for placebo arm A-CDP predictor are displayed. **E.** Relationship between baseline ARR and efficacy of MS DMTs on A-CDP (i.e., A-CDP%Δ with smaller negative values representing higher efficacy) in placebo-controlled clinical trials. The R^2^ of A-CDP%Δ predicting model and p-values of its predictors (ARR and Trial/treatment duration coded as color with short duration in blue and long in orange) are displayed. ARR increases efficacy whereas treatment duration decreases efficacy. **F.** Panel analogous to panel D, except now showing also active comparator trials (highlighted with red outer circle) for which we computed A-CDP in the “in-silico placebo arm”, based on predicted efficacy of any DMT on recruited patient population modified by mean weighted residual of the utilized active comparator as described in Methods and Supplementary information. Including active comparator trials did not significantly change the relationship between A-CDP in (real or in-silico predicted) placebo and active treatment arms. **G.** Comparative efficacy of MS drugs based on the residuals between Eq#6-predicted and measured efficacies on A-CDP. MS DMTs are arranged from top (orange colors; high efficacy) to bottom (blue colors low efficacy) based on decreasing comparative efficacy. Small trials (weight <700; omitted from comparative efficacy calculations) are displayed in gray, due to their outlier status. Trials codes can be linked to specific trials listed in the “index” column of the SSW Master Worksheet. We highlighted in blue font drugs that were taken off marked for toxicity (daclizumab) or never achieved FDA approval (laquinimod and high dose biotin) while in pink are drugs with discordant residuals for MS type (natalizumab) or drug dose (ozanimod, although trial design pooled both doses together).

As ARR decreases with time (Figure 3), *Eq#4* implies two *opposite* contributors to A-CDP: LA (reflected by ARR or CEL#) rapidly decreasing after MS onset and progression independent of LA (PILA) steadily increasing with DD (Figure 4B).

Supporting two separate A-CDP contributors, we observed that when using all trial arms (i.e., placebo + DMT; Figure 4C), on-trial ARR explains 54% of A-CDP variance in RRMS patients with DD having less effect, whereas including SPMS trials decreased ARR contribution to 22% with DD explaining most residual variance.

### Baseline patient characteristics predict A-CDP better than treatment status

Next, we correlated A-CDP rates between the placebo and treatment arms (Figure 4D). Strong correlation (Rho = 0.94, p<0.0001) indicates that baseline characteristics of recruited populations are dominant determinants of A-CDP in treated patients. Consistently, the multivariate predictor of treatment A-CDP [*Eq#5*] explains 91% of variance. In addition to placebo-arm progression rates (p<0.0001), the model includes EDSS (p=0.0015) and LA captured by the inclusion criterion of minimum CEL# (p=0.12; *SSW-s9*).

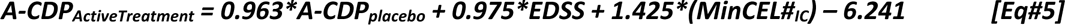

*Eq#4* shows placebo arm A-CDP depends on ARR and DD; therefore, the baseline patient characteristics (i.e., ARR, CEL#, DD, EDSS) predict A-CDP rates much stronger than treatment status.

### DMT efficacy on A-CDP increases with LA and decreases with DD and treatment duration

Efficacy on A-CDP is calculated as the relative difference in the proportion of patients progressing yearly on treatment versus placebo (i.e., A-CDP%Δ), with higher efficacy corresponding to *negative* A-CDP%Δ.

We sought to predict DMT efficacy on A-CDP with LA markers (I/E criteria of Minimum ARR and Minimum CEL#, baseline mean CEL# and ARR), time-related predictors (trial duration, DD, age), EDSS and sex. The final model ([*Eq#6*], *SSW-s10*) explains 75% of variance and shows that efficacy decreases with trial duration (p=0.0166) and DD (p=0.1268) and increases in patients with higher ARR (p=0.0346; Figure 4E):

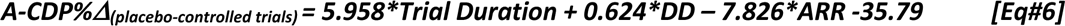

We conclude that from MS disability progression determinants (Figure 4B) current DMTs limit only disability accumulation from LA (measured by ARR); DMT efficacy decreases with DD, which reflects PILA. Furthermore, the ability of DMTs to inhibit A-CDP decreases after treatment initiation with unexpectedly strong effect size: each treatment year decreases efficacy by ∼6% (95% CI = 1.247-10.669).

### Baseline characteristics of trial population determine dynamic aspect of treatment effect

The 6% represents absolute yearly efficacy decline: e.g., in cohorts with 50% predicted efficacy, DMTs will continuously slow disability accumulation for ∼8.3 years, whereas inhibition of A-CDP is predicted to last ∼3.3 years in patients with 20% predicted efficacy.

Consequently, we hypothesized and observed that the short-lasting treatment effects that *Eq#6* predicts for older, more disabled pwMS are discernable within trial duration; indeed, the relevant cohort characteristics (i.e. LA and DD) determined both static efficacy on A-CDP and its dynamic aspect observed in published Kaplan-Meier (K-M) survival curves.

The dynamic aspect of K-M curves contains valuable information about DMT’s mechanism of action and predicts DMTs efficacy beyond trial duration. In cohorts with high LA and low DD, K-M curves showed sustained A-CDP%Δ (Supplementary Figure 5A; e.g., IFN-β RRMS^4, 5^ trials and ocrelizumab OPERA I/II trials^6^). We conclude that in these early RRMS patients therapeutic effect starts at treatment initiation and continues for the trial duration. Conversely, in cohorts with high DD and low LA, K-M curves showed declining efficacy (e.g., IFN-β SPMS trials^7, 8^ and ocrelizumab PPMS ORATORIO^9^ trial), where treatment and placebo arms maximally separated in the 1^st^ year, after which A-CDP rates in both arms remained identical. Thus, in these pwMS DMTs produced immediate therapeutic effect that wasn’t sustained, because DMTs inhibited disability caused by LA without affecting disability progression due to PILA. Cumulatively, this resulted in *delaying* rather than *eliminating* disability progression.

Final K-M curve dynamic pattern not observed in MS trials (Supplementary Figure 5A, right panel) is *increasing* efficacy with treatment duration. Because PILA mechanisms’ effect on disability progression increases with time (Figure 4A,B), the therapeutic effect of PILA-inhibiting DMTs should also increase with treatment duration. The absence of this K-M pattern strengthens our conclusion that current DMTs inhibit LA but not PILA mechanisms of disability accumulation.

K-M curve dynamics also strengthen projections of therapeutic efficacy beyond trial duration (i.e., static, increasing or decreasing). Therefore, we digitalized the published curves and derived a Time Delay variable (TD; Supplementary Figure 5B; Supplementary methods). TD ranges from 0-1; 0 represents no yearly delay in disability progression by DMT, 0.5 represents a 6-month delay and 1 represents a 12-month delay. A-CDP%Δ and yearly TD from placebo-controlled trials correlated weakly (R^2^ = 0.386, p=0.0016), demonstrating that TD provides non-redundant information. Assuming that for most pwMS long-term DMT administration only delays disability progression, we used TD as a modifier of static efficacies: the resulting novel outcome (A-CDP*TD%Δ) integrates both static and dynamic aspects of K-M curves. Expectedly, A-CDP%Δ and A-CDP*TD%Δ correlated (Supplementary Figure 5C; weighted r_Pearson_ = 0.834, R^2^=0.696, p<0.0001) but A-CDP*TD%Δ showed a lower, more realistic estimate of MS DMTs long-term efficacies.

### Comparative efficacies of MS DMTs

Although baseline characteristics of the recruited population explained 91% of A-CDP rates of treated cohorts (Figure 4D), the remaining 9% could be due to differences in efficacies among MS DMTs. We aimed to compare efficacies between DMTs adjusting for the baseline characteristics of recruited populations. However, we had to first re-calculate efficacies for trials that used an active comparator instead of a placebo.

Twenty-four clinical trials used an active comparator but only 19 reported CDP. A candidate drug is considered superior when significantly fewer candidate drug-treated patients progress by trial end compared to patients on an older comparator. However, trial design may invalidate this interpretation. We identified and corrected for design biases that disadvantage comparators (Supplementary methods) and used *Eq#4&6* to predict A-CDP in “in-silico placebo arms” (Figure 4F).

Residuals’ variance from observed versus *Eq#6*-predicted A-CDP%Δ (Figure 4G) implied that for some DMTs the same drug showed dissimilar efficacies. Although this may reflect differential effects of the drug on LA vs PILA biology (likely for natalizumab), or of different drug doses (e.g., ozanimod trials), small trial imprecision explained a large part of this variance (Supplementary Figure 6). Thus, we excluded small trials (i.e., A1, D1, F5, I1, M1, R2 and T3) from comparative analyses.

Of the remaining trials, B cell depleting treatments ocrelizumab and ofatumumab, and alemtuzumab (depletes B cells together with other immune cells) demonstrated the highest efficacies. However, rater-blinded only design may have overestimated alemtuzumab’s efficacy.

Moderately effective drugs are selective S1PR1 modulators ponesimod and siponimod (binds S1PR1&5), daclizumab (taken off the market) and fumarate-based preparations.

Drugs with efficacy close to *Eq#6* prediction are all interferon-β and glatiramer acetate preparations, non-selective S1PR modulator fingolimod, chimeric B cell-depleting antibodies rituximab and ublituximab, and (not FDA approved) laquinimod.

Teriflunomide and ozanimod have below-average efficacy. RRMS natalizumab trial (N1) has moderate efficacy while its PMS trial (N3) may have been harmful.

### DMTs increase risk of infections

Because DMTs target the immune system, we assessed risks of infections and cancer.

Rare, serious infectious complications of MS DMTs^10, 11^ are partially mitigated by surveillance, vaccinations and preventive strategies^12^: progressive multifocal encephalopathy (natalizumab, fingolimod, dimethyl fumarate), cryptococcal meningitis (fingolimod), tuberculosis reactivation (natalizumab, fingolimod, mitoxantrone, teriflunomide), herpes simplex/zoster reactivation (alemtuzumab, fingolimod), hepatitis B reactivation (B cell-depleting drugs, alemtuzumab, mitoxantrone).

Additionally, DMTs effects on common infections cause higher population morbidity/mortality. Congruent population studies (Supplementary results) show DMTs, age, high disability and comorbidities increase risk of infections^13,14, 15^. Assuming that severe infection mortality is identical between MS and the general population, we superimposed adjusted risk ratios^13^ for severe infections onto Center for Disease Control and Prevention (CDC) “all infections” mortality tables (Figure 5A; SSWs12-14). MS patients have increased morbidity/mortality from infections compared to the general public, which is further increased by DMTs (Figure 5B). Indeed, MS patients older than 45 years have an 80% higher overall mortality^16^ than the general public (Figure 5C), decreasing their life expectancy (Figure 5D). By increasing infections, MS DMTs are responsible for part of observed mortality rise.

**Figure 5:**
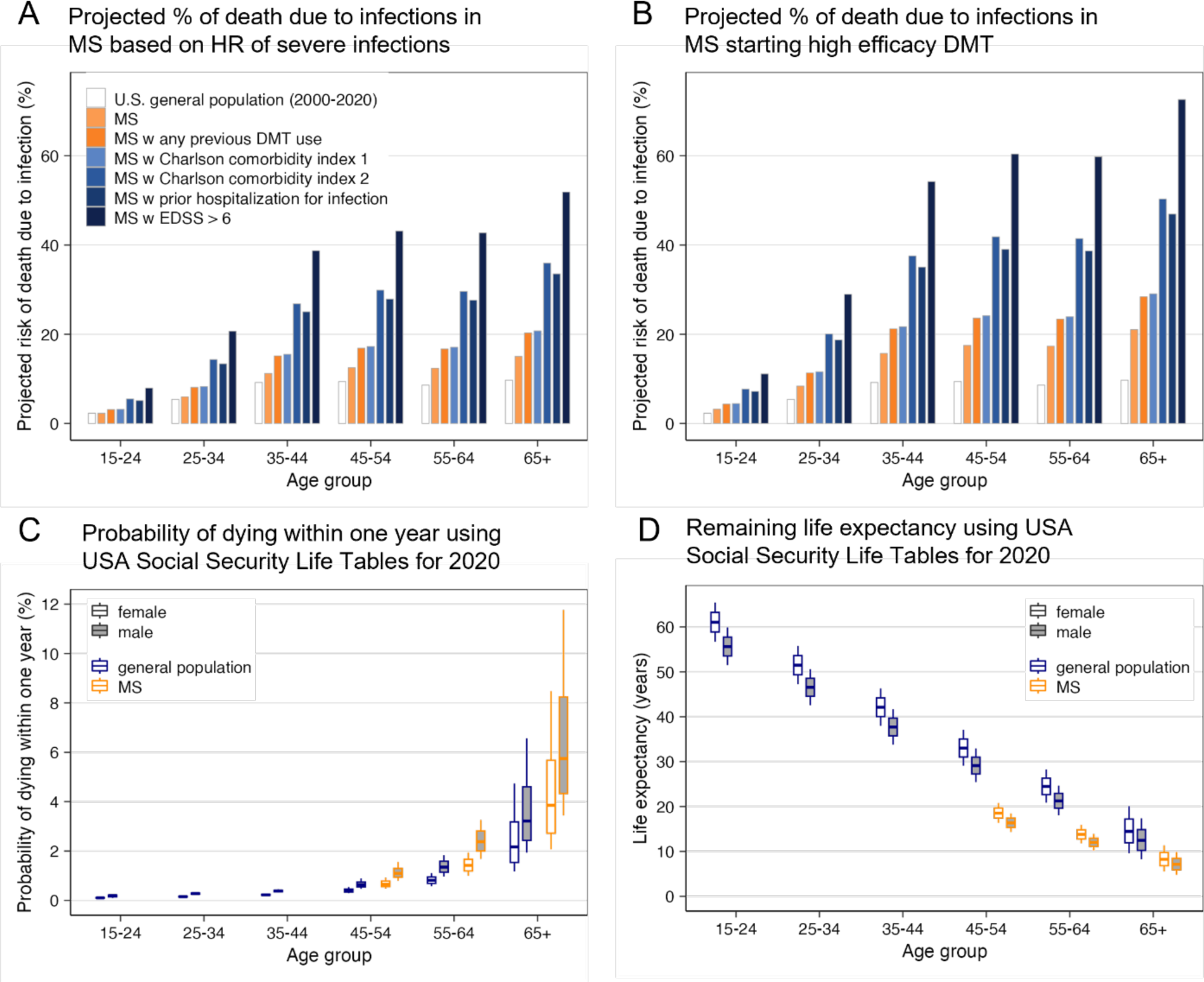
MS patients have increased mortality and decreased life expectancy, partially driven by age-, disability- and comorbidities-related increased risk of severe infections. High efficacy DMTs further enhance infectious morbidity/mortality. **A.** Bar chart of age-related risk of death due to infections. The empty bars represent proportion of deaths due to any type of infections reported in USA CDC mortality 2001-2020 tables. Assuming this represents general population relative risk of infectious death, and further assuming that mortality from severe infections is equal between MS and general population, we super-imposed published adjusted Hazard Ratios (aHR) for severe infections in USA MS population to estimate risk of infection-related mortality for MS pations: the light orange bars show that compared to general population, MS patients have age-related increase in incidence of severe infections, which is further increased in MS patients with previous history of treatment with any MS DMT (dark orange bars). Risk of severe infections is further increased in MS patients with comorbidities measured by Charlson Comorbidity Index (CCI; two lightest colored blue bars). Finally, MS patients with prior hospitalization for infections (5^th^ bar, second darkest blue) have similar risk of death from infections to patients with CCI ≥2, but the highest risk of infectious mortality is for patients with EDSS>6, who are either non-ambulatory or require bilateral assistance to ambulate. **B.** Identical data to panel A but modified by aHR = 1.4 of MS DMTs. We note that from DMTs with available data, this aHR reflects only rituximab, natalizumab and fingolimod and excludes interferon-beta and glatiramer acetate (GA) preparations that do not increase risk of severe infections. **C.** Yearly probability of death due to all causes derived from USA Social Security 2020 Period Life Table (https://www.ssa.gov/oact/STATS/table4c6_2020_TR2023.html; general population blue outline) separated for males (grey filled box plots) and females (empty box plots) and modified by published aHR = 1.8 of MS risk of death^16^ for patients 45-79 year old (orange outline; no data available for younger patients). The box plots show median (horizontal line), interquartile range and minimum/maximum values for subjects in each age decade. **D.** Remaining life expectancy derived from USA Social Security 2020 Life tables for general population (blue outline) modified by published aHR = 1.8 of MS-related risk of death^16^ (0.55 probability of survival) for patients aged 45-79 years. The box plots show median (horizontal line), interquartile range and minimum/maximum values for subjects in each age decade.

Unfortunately, we found insufficient data on DMTs’ risks of cancer (Supplementary results). While package inserts state that ocrelizumab increases risk of breast cancers, fingolimod skin cancers and mitoxantrone acute myeloid leukemia, population studies show that <4 years administration of (older) DMTs to cohorts with mean age ≤ 40 years does not increase de-novo cancer incidence^17, 18^. In contrast, the World Health Organization VigiBase® suggests that long-term administration of MS DMTs increases cancer risk with odd ratios between 1.15-1.74^19^.

We conclude that a lack of studies prevents estimating how DMTs affect incidence of primary and secondary cancers and cancer severity. As cancer mortality exceeds infectious mortality ∼3-fold, even a marginal increase in cancer incidence or severity would substantially increase MS mortality.

### Web-based estimator of patient-specific DMT risk/benefit ratios

We aimed to translate clinical trial’ insights and real-world MS DMT risk data to patient-specific risk/benefit estimates. Because real-world predictors of DMTs’ efficacy don’t exist, extrapolating clinical trial equations to the general MS population requires careful consideration.

Predictor values (ARR, CEL#, age, EDSS, DD) vary more in MS population than in trial-recruited patients. This artificial homogeneity among predictor averages in trial populations underestimates the effects that predictors not selected by stepwise models likely play in the diverse MS population. For example, Figures 4D&F predict that among pwMS of the same age, ARR and DD, mildly disabled patients will benefit more from DMTs than severely disabled patients, even though EDSS was not retained in multivariate efficacy model, likely due to collinearity with DD. Furthermore, personalized efficacy estimates cannot be generated from multivariate models for people lacking (some) required predictors.

To mitigate these limitations we generated simple weighted A-CDP%Δ correlations for efficacy predictors available in clinical practice (Figure 6A; SSWs11). We generated analogous correlations for the more realistic, A-CDP*TD%Δ outcome, which assumes that MS drugs delay, not abrogate disability progression for most pwMS. Though we could compute A-CDP*TD%Δ only for 29 trials, it correlated comparably or stronger with efficacy predictors (Figure 6B; SSWs11). Finally, while we avoided active-comparator trials for modeling due to multiple imputations, their inclusion does not alter efficacy estimates (Figure 6C vs. Figure 6A).

**Figure 6:**
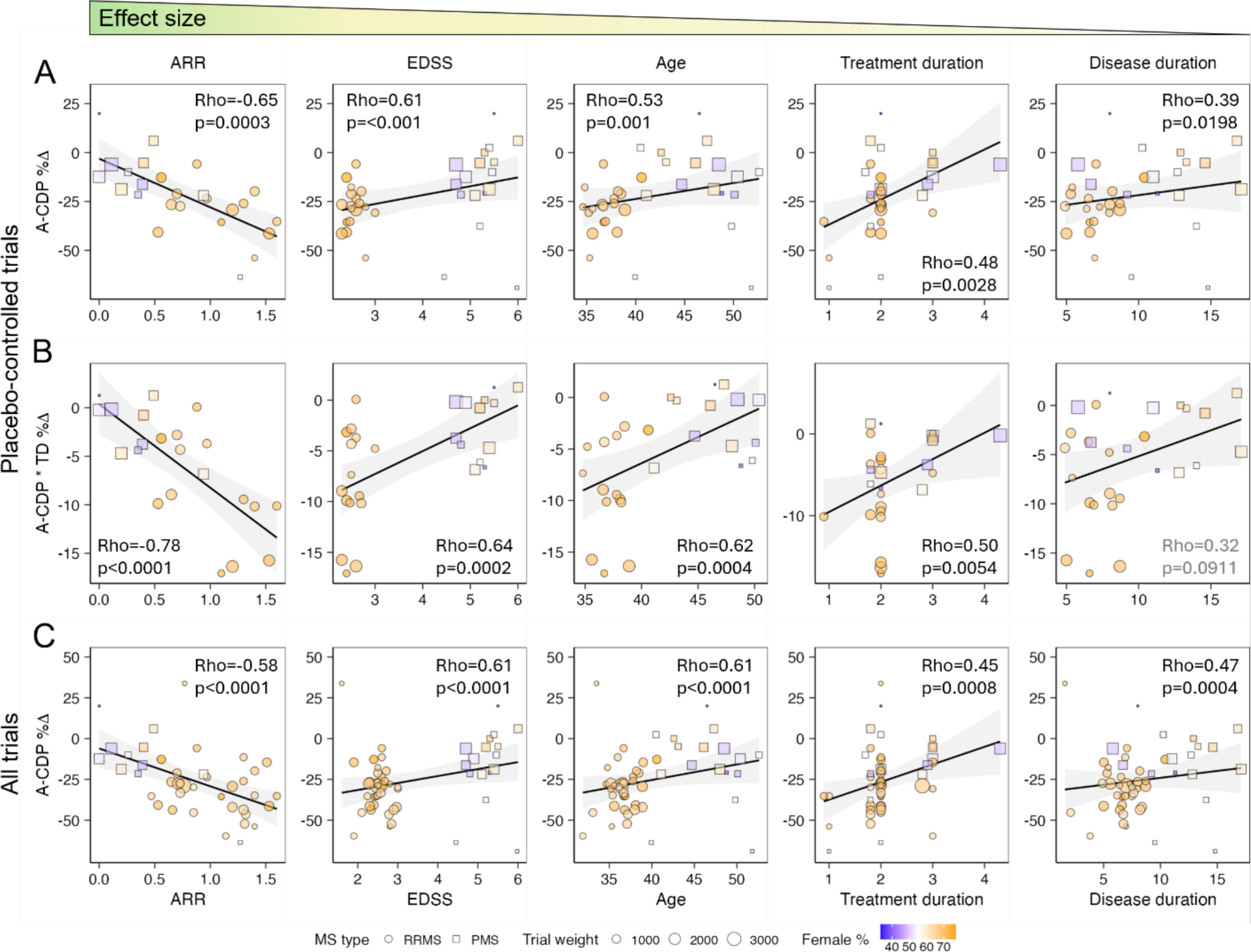
Modeling DMT efficacy from group means under-estimates effects of diverse disease characteristics on disability progression, and over-estimates efficacy by ignoring dynamic aspect of the therapeutic effect (i.e., MS DMTs on average only delay, rather than prevent disability progression) **A.** Simple weighted correlations of efficacy predictors of A-CDP%Δ from placebo-controlled trials, arranged from left to right with decreasing effect sizes. Pearson Rho and p-values for each predictor are displayed. For all panels, RRMS trials are shown as circles and PMS trials as squares with size proportional to trial weight 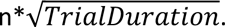 Proportion of females in trial population is coded according to color heatmap (fewer females in blue, more in orange). **B.** Analogous plots of efficacy predictors with novel efficacy outcome that incorporates time-delay (TD) variable from published Kaplan-Meier survival curves (i.e., A-CDP*TD%Δ). This outcome does not assume that MS DMTs prevent disability progression; instead TD efficacy modifier measured over first 2 years of each clinical trial that lasted ≥2 years and published Kaplan-Meier curves reflects the amount of time by which DMT delayed sustained disability progression. Because none of MS DMTs are curative, this is more realistic estimate of DMT efficacy for most patients. Shown p-values are expected to be weaker because of smaller number of trials for which this outcome could be calculated. **C.** Panel analogous to A (i.e., showing weighted Pearson correlations with traditional A-CDP%Δ outcome, but this time including active comparator trials. Including active comparator trials does not substantially change effect sizes of efficacy predictors (p values are stronger due to larger number of trials).

We implemented predictions for both efficacy outcomes in a web-based tool https://bielekovalab.shinyapps.io/shinyapp/ that estimates DMTs efficacy, requiring the patient’s age at minimum. We provide weighted-average efficacy estimate when more predictors are submitted. In addition to estimating *average* efficacy of DMTs based on patient characteristics (Figure 7A), selecting a specific DMT from the drop-down menu applies Figure 4G comparative efficacy to generate personalized, drug-specific efficacy estimates.

**Figure 7:**
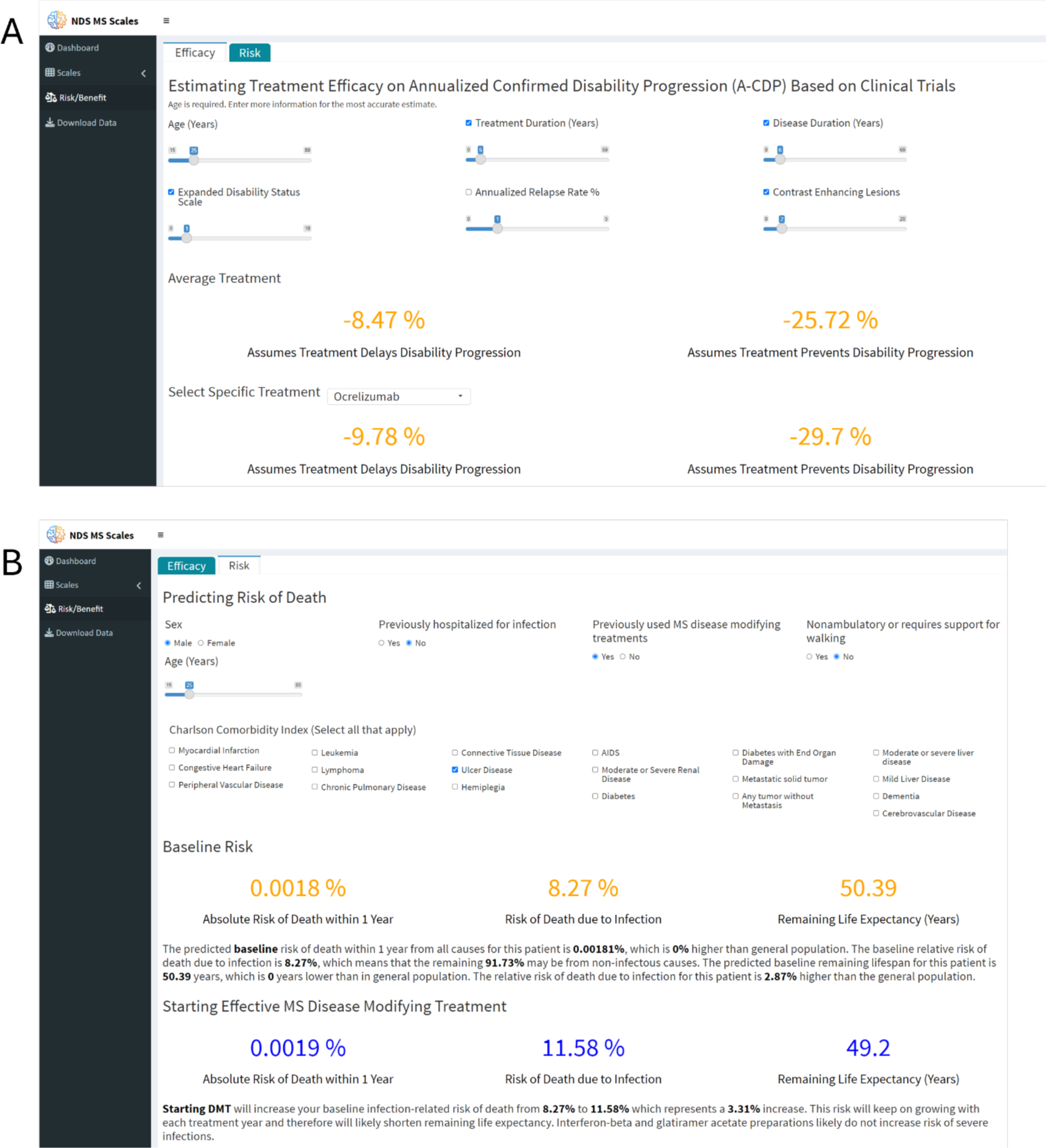
Interactive website for calculating patient-specific risk/benefit predictions. The risk/benefit predicting tool is generated in ShinyApp by R studio and hosted together with previously developed digital tools from Bielekova laboratory: https://bielekovalab.shinyapps.io/shinyapp/. To navigate to Risk/benefit prediction, user selects appropriate Tab on the left side menu. Risk/benefit tool has two Tabs: **A:** Efficacy prediction requires minimum of Age input, but selecting more efficacy predictors provides more accurate (weighted) average. The efficacies for more realistic A-CDP*TD%Δ outcome that assumes that DMT delays disability progression (displayed on left side) and traditional A-CDP%Δ that assumes that DMT prevents disability progression (displayed on right side of the screen) change dynamically whenever user imputes/changes predictor value(s). Changing treatment duration slider then allows predicting efficacy during the proposed length of treatment. **B.** Risk predictor tab first calculates baseline risk of infectious mortality, overall mortality and predicted remaining lifespan for specific patient. Again, risk could be calculated from age only, but answering remaining questions about previous hospitalization for infections, severe disability, history of previous DMTs and clicking existing comorbidities to calculate Charlson Comorbidity Index prevised more accurate estimates. After selecting new DMT, the website then compares the existing baseline risk of infection-related and overall mortality/lifespan with new predicted risks that include effect of high efficacy DMTs on these baseline predictions.

The second web-based tool estimates DMTs risks (Figure 7B). The baseline and DMT-mediated morbidity can be estimated from age only, but selecting risk modifiers such as previous DMT use, previous hospitalizations for infections, disability and comorbidities (to calculate Charlson Comorbidity Index^20^) strengthens risk estimates.

The website interactively modifies risk/benefit estimates with treatment duration, informing treatment initiation and optimal time to treatment de-escalation or discontinuation.

## Discussion

By providing accessible, data-driven estimates of risk/benefit for DMTs based on individual patient characteristics, this study advances MS treatment optimization and may save countless iatrogenic hospitalizations and deaths.

Study limitations (beyond authors’ control) include lack of patient-level data and real-word DMT efficacy data as well as estimates of DMTs’ long-term cancer risks. Lack of access to raw data from clinical trials^21^ could be remedied if publishers required reporting descriptive statistics for predictors and outcomes for patients in each age decade. Assessing long-term infectious and cancer risks of immune-targeting DMT should become a healthcare priority. Finally, real-world efficacy data may come from Patient-Centered Outcomes Research Institute sponsored trials.

One of these trials, the DISCOMS^22^ already demonstrated that randomized discontinuation of DMTs in pwMS (median age 63y, 83% RRMS, mean EDSS 3.4) who lacked LA for at least 3 years during treatment did not increase CDP rates over the subsequent 24 months. At trial conclusion pwMS who discontinued DMTs had insignificantly improved mean EDSS (-0.1) than pwMS who continued DMTs (+0.1 EDSS change). However, more pwMS who discontinued DMTs experienced LA (2.3% experienced relapse and 10.7% experienced new T2 lesion) than pwMS who continued treatment (0.8% with relapse and 3.9% with new lesion). Consistent with our models, pwMS experiencing LA after DMT discontinuation were slightly younger, less disabled, more likely to have RRMS, and more likely treated with higher-efficacy DMTs; however, numbers were too small to achieve statistical significance.

In conclusion, DMT discontinuation can lead to re-appearance of LA without impacting (short-term) disability progression rates. PwMS in DISCOMS were not preselected for DMT discontinuation based on risk/benefit estimates defined in this study. Additionally, LA re-appearance would increase predicted DMT efficacy in our estimator. This should prompt new discussion between the prescriber and patient about the risk/benefit of reinstating DMT. Monitoring for LA reappearance after DMT discontinuation can be economically achieved also by serum neurofilament light chain (NFL)^23, 24^.

Apart from developing personalized risk/benefit estimator, our study provides holistic, unified view of MS evolution and identified or validated following insights:

Over 3 decades of drug development, trial patients have progressively less severe MS, which may favor comparative efficacy of newer drugs. While the enrollment of partially treated patients and the trial avoidance of subjects with active MS are attractive explanations for declining MS severity, similar declines were observed in MS population studies after adjusting for treatments^25^. Therefore, this severity decline may be caused by shifting emphasis from clinical to radiological criteria in MS diagnosis and public health initiatives such as smoking cessation and vitamin D supplementation^25^.

Our models show that current DMTs inhibit disability progression caused by LA but are ineffective against PILA-related disease mechanisms. We deliberately avoided the previously proposed term “Progression Independent of Relapse Activity (PIRA)”, as PILA more accurately identifies patients who do not benefit from current DMTs. Because not every CEL causes an MS relapse, patients fulfilling PIRA may still form CELs, such as PPMS patients recruited to the OLYMPUS^26^ and ORATORIO^9^ trials. To the extent to which pwMS fulfilling PIRA criteria form CELs, our results predict that they will benefit from current DMTs. This is what both PPMS trials observed. In the ORATORIO trial, 27.5% of PPMS patients randomized to ocrelizumab and 24.7% randomized to placebo had CELs at baseline. As ocrelizumab rapidly inhibits CELs^6^, ocrelizumab should equally rapidly inhibit the proportion of A-CDP caused by LA and generate early separation of K-M curves. Indeed, ocrelizumab and placebo arms separated within 12 weeks. Because ocrelizumab almost completely^6^ inhibits CEL#, ocrelizumab-treated patients must have progressed by PILA mechanisms after 12 weeks. And after week 12, both ocrelizumab and placebo arms progressed linearly and in parallel to each other, indicating that ocrelizumab has no additional efficacy on PILA. Consequently, PILA-targeting treatments are greatly needed. Our models predict that efficacy of such treatments should increase with treatment duration, suggesting that PILA-targeting trials have to be sufficiently long.

Previous meta-analysis^2^ showed that DMT efficacy decreases with increasing age at treatment onset. This study provides new insight that DMT efficacies decline rapidly *with treatment duration*. Delay between MS onset and DMT initiation reduces life-long efficacy that cannot be recovered. Practical screening that shortens diagnostic delay (e.g., identifying people with neuro-axonal injury using NFL and referring them for diagnostic work-up) would have strong population-level benefit. While high efficacy DMTs initiated at MS onset maximizes efficacy, their subsequent de-escalation to DMTs that do not increase infectious morbidity (glatiramer acetate, IFN-β) would minimize risk. Personalized risk/benefit estimates can guide the timing of such DMT de-escalation or discontinuation.

Presented results show that most people living with MS today^27^ (i.e., older than 55y, disabled, lacking LA) do not benefit from current DMTs. If treated, they have an increased infection- (and likely cancer)- related morbidity and mortality. Making treatment decision based on data-driven risk/benefit estimates will optimize MS care while financial savings can be redirected to non-pharmacological interventions, such as physical and occupational therapy.

## Supporting information

Supplementary informtion

URL for analyzed clinical trials

## Abbreviations and glossary terms

A-CDP: Annualized confirmed disability progression (proportion of subjects reaching A-CDP per each year of trial)
A-CDP%Δ: Efficacy on the annualized confirmed disability progression – because it measures difference in A-CDP between treated and untreated subject(s), higher efficacy is reflected by lower numbers
A-CDP%Δ*TD: Efficacy on annualized confirmed disability progression modified by time delay (TD) variable. Unless the drug prevents disability progression in all treated patients, this efficacy is always smaller than A-CDP%Δ because it does not assume that people who did not progress during trial duration will never progress; instead it assumed that MS drugs on average only delay, rather than completely prevent disability progression. TD variable computes the amount of this delay from published Kaplan-Meier survival curves. This is more realistic efficacy outcome than A-CDP%Δ
ARR: Annualized relapse rate (number of relapses per year)
CEL#: Number of contrast enhancing MS lesions on brain MRI
DD: MS duration. Calculated in years from first MS symptom
DMTs: disease modifying therapies
LA: Lesional activity. This term encompasses formation of new MS lesions, measured as contrast-enhancing lesions (CEL) or new T2 lesions and clinically represented by MS relapses
MS: multiple sclerosis
PILA: Progression independent of MS LA. Identifies patients with sustained disability progression who neither experience relapses nor form new or contrast enhancing MS lesions
PIRA: Progression independent of relapse activity. Identifies patients with sustained disability progression without MS relapses. These people may still have contrast enhancing lesions on brain and spinal cord MRI
PPMS: Primary progressive MS. Patients who never experienced MS relapses and are progressing. They can form contrast enhancing lesions on MRI of brain or spinal cord
Progressive MS: all patient who are progressing outside of relapse activity (PPMS+SPMS)
RRMS: Relapsing remitting MS. Patients who are experiencing MS relapses
Relapse onset MS: all MS patients who experience or experienced MS relapse (RRMS + SPMS)
Residual variance: statistical term that reflects imprecision of the model’s prediction. If model predicts outcome with 100% accuracy, the residual variance is zero. Residual variance may represent noise, or the proportion of the outcome that is determined by the predictor that is not available (e.g., unknown or not measured).
SPMS: Secondary progressive MS. Patients who experience relapses or experienced relapses at MS onset, but are now progressing between relapses or without relapses
Stepwise multiple regression model: statistical prediction of continuous outcome (such as probability of annualized confirmed disability progression) from multiple predictors. Selection of predictors occurs in stepwise fashion and is guided by statistical significance that reflects the probability that including the predictor makes outcome prediction meaningfully stronger
SSW: Supplementary Statistical Workbook
TD: time delay variable derived from published Kaplan-Meier survival curves. It reflects the yearly delay of disability progression and varies from 0-1, with 0 representing no delay and 1 representing full year.

## Funding

This work has been supported by the intramural research programs of the National Institute of Allergy and Infectious Diseases (NIAID) and National Institute of Neurological Disorders and Stroke (NINDS).

## Acknowledgements

We thank Drs. Annette M. Langer-Gould and Gary Birnbaum for critical review of the manuscript and helpful comments. This paper is dedicated to the memory of Henry McFarland, a devoted MS physician-scientist, leader and mentor who left us way too early.

## Author Contributions

BB contributed to conception and design of the study; BB and TW contributed to the acquisition and analysis of data; BB, TW, MC, and PK contributed to drafting of the text or preparing the figures.

## Potential Conflicts of Interest

Nothing to report.

## Data availability

The data that supports the findings of this study are available in the supplementary material of this article.

## Notes

### Competing Interest Statement

The authors have declared no competing interest.

### Author Declarations

The study used only openly available human data there were originally published in Pubmed - URL for the studies are available in supplementary files

