## Supplementary informtion for "Data-driven risk/benefit estimator for multiple sclerosis therapies"

### Supplementary Appendix

#### Abbreviations and glossary terms

A-CDP: Annualized confirmed disability progression (proportion of subjects reaching A-CDP per each year of trial)

A-CDP%∆: Efficacy on the annualized confirmed disability progression – because it measures difference in A-CDP between treated and untreated subject(s), higher efficacy is reflected by lower numbers

A-CDP%Δ*TD: Efficacy on annualized confirmed disability progression modified by time delay (TD) variable. Unless the drug prevents disability progression in all treated patients, this efficacy is always smaller than A-CDP%Δ because it does not assume that people who did not progress during trial duration will never progress; instead it assumed that MS drugs on average only delay, rather than completely prevent disability progression. TD variable computes the amount of this delay from published Kaplan-Meier survival curves. This is more realistic efficacy outcome than A-CDP%Δ

ARR: Annualized relapse rate (number of relapses per year)

CEL#: Number of contrast-enhancing MS lesions on brain MRI

DD: MS duration. Calculated in years from first MS symptom

DMTs: disease modifying therapies

LA: Lesional activity. This term encompasses formation of new MS lesions, measured as contrast-enhancing lesions (CEL) and clinically represented by MS relapses

MS: multiple sclerosis

PILA: Progression independent of MS LA. Identifies patients with sustained disability progression who neither experience relapses nor form new or contrast enhancing MS lesions

PIRA: Progression independent of relapse activity. Identifies patients with sustained disability progression without MS relapses. These people may still have contrast enhancing lesions on brain and spinal cord MRI

PPMS: Primary progressive MS. Patients who never experienced MS relapses and are progressing. They can form contrast enhancing lesions on MRI of brain or spinal cord

Progressive MS: all patient who are progressing outside of relapse activity (PPMS+SPMS)

RRMS: Relapsing remitting MS. Patients who are experiencing MS relapses

Relapse onset MS: all MS patients who experience or experienced MS relapse (RRMS + SPMS)

Residual variance: statistical term that reflects imprecision of the model's prediction. If model predicts outcome with 100% accuracy, the residual variance is zero. Residual variance may represent noise, or the proportion of the outcome that is determined by the predictor that is not available (e.g., unknown or not measured).

SPMS: Secondary progressive MS. Patients who experience relapses or experienced relapses at MS onset, but are now progressing between relapses or without relapses

Stepwise multiple regression model: statistical prediction of continuous outcome (such as probability of annualized confirmed disability progression) from multiple predictors. Selection of predictors occurs in stepwise fashion and is guided by statistical significance that reflects the probability that including the predictor makes outcome prediction meaningfully stronger

SSW: Supplementary Statistical Workbook

TD: time delay variable derived from published Kaplan-Meier survival curves. It reflects the yearly delay of disability progression and varies from 0-1, with 0 representing no delay and 1 representing full year.

### Supplementary Methods

#### Supplementary Figure 1: PRISM diagram

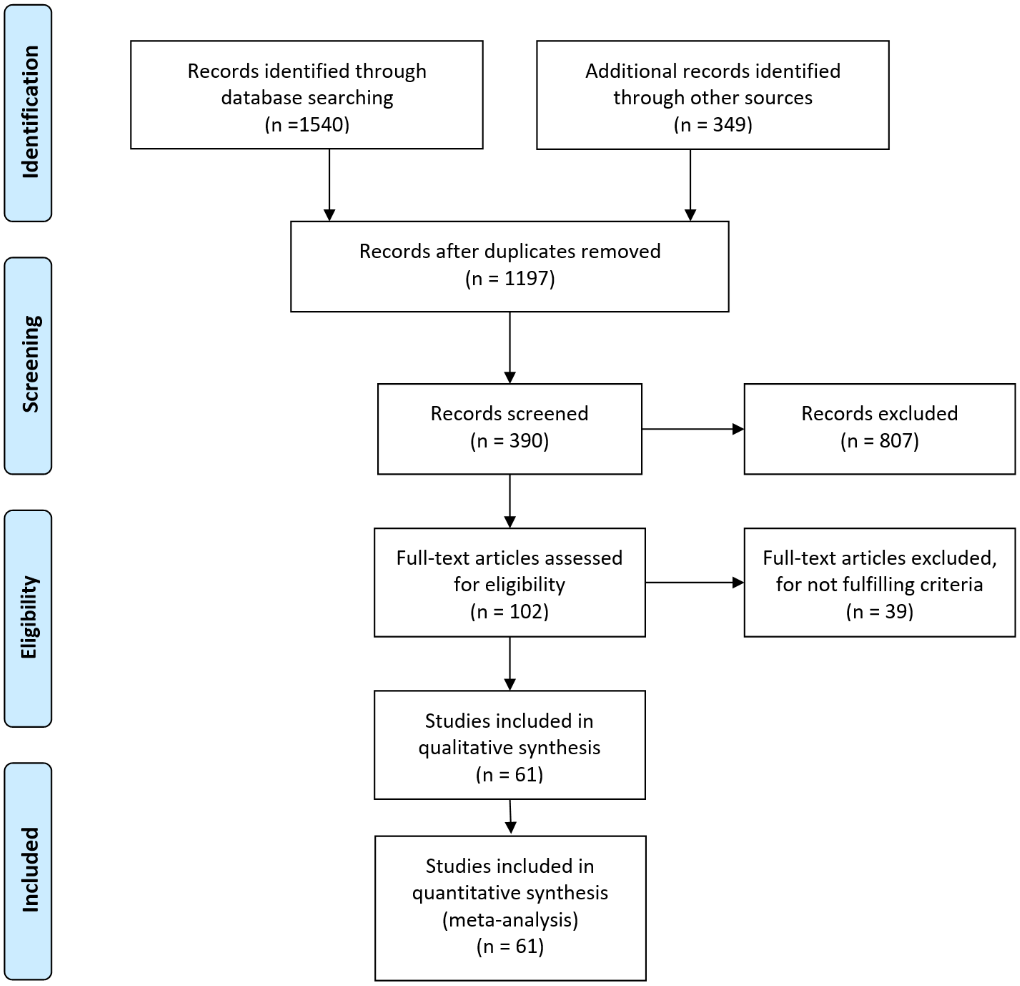

#### Categories of extracted and imputed data elements

For both active treatments and control arms, we systematically extracted 80 data elements (Supplementary Statistical Workbook [SSW], Master tab; categories described below):

A. Trial design metadata: 16
B. Inclusion/Exclusion criteria: 10
C. Baseline characteristics of the recruited population: 23
    a. Demographic: 5
    b. Clinical: 8
    c. MRI: 10
D. On trial measurements: 31
    a. Clinical: 10

b. MRI: 21

Additionally, after identifying statistically significant relationships of moderate/high effect sizes (i.e., $R^2$>0.5) in the extracted data, we computed 30 additional features:

A. Trial-related: 2
B. Baseline characteristics (MRI based): 2
C. On-trial measurements: 26
   a. Clinical: 16
   b. Clinical derived from digitalization of published Kaplan-Meier curves: 10

#### Supplementary Figure 2: Imputation of CEL# (average number of CELs/scan) for studies that published only CEL% (proportion of patients with CELs)

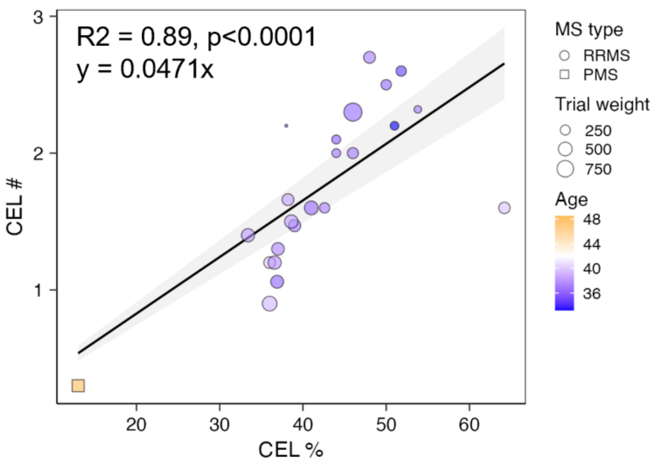

**Supplementary Figure 2**

Relationship between proportion of patients with CELs (CEL%) and average number of CELs/MRI (CEL#) from trials that reported both parameters. RRMS trials are shown as circle, progressive MS trials as squares, with size proportional to trial weight. Mean age of recruited population is displayed as heatmap color.

#### Supplementary Figure 3: Imputation of confirmed disability progression (CDP) confirmed at 12 weeks (CDP@12wk) for trials that only reported CDP confirmed at 24 weeks (CDP@24wk)

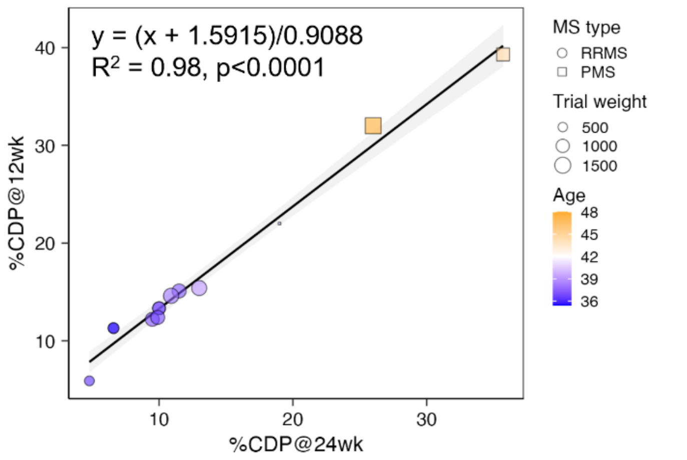

**Supplementary Figure 3**

Relationship between proportion of patients achieving disability progression that was confirmed at 12 weeks (%CDP@12wk) and also at 24 weeks (%CDP@24wk) from trials that reported both. RRMS trials are shown as circle, progressive MS trials as squares, with size proportional to trial weight. Mean age of recruited population is displayed as heatmap color.

#### Supplementary Figure 4: Imputation of DD using baseline patient characteristics for trials that did not report DD

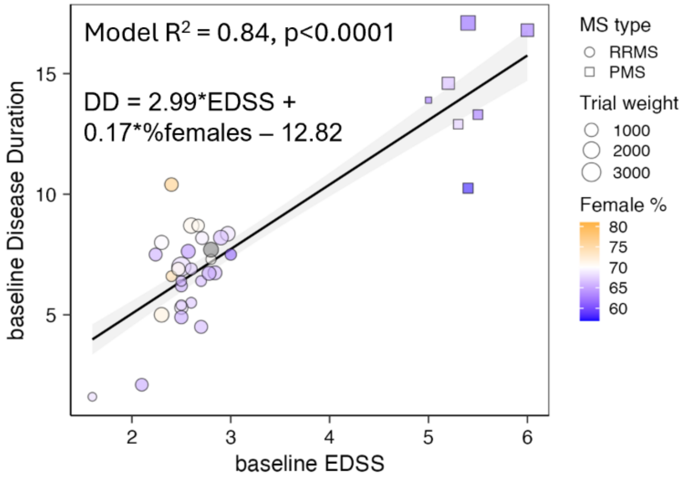

**Supplementary Figure 4**

Depiction of the relationship between baseline EDSS, disease duration (DD) and proportion of females in recruited population (%females) from trials that reported all parameters that served as basis for stepwise multiple regression model used for DD imputation. The model $R^2$, p-value of EDSS predictor and the equation used for DD imputation using EDSS and %females is depicted in the Figure. RRMS trials are represented with circles, progressive MS trials with squares (with size proportional to trial weight). % of females in the recruited population is displayed as heatmap color.

#### Analyses of dynamic data from Kaplan Meier (KP) survival curves

The dynamic analysis of the treatment effect stems from the hypothesis that MS DMTs only *delay* disability progression in most patients (rather than *preventing* disability progression in its entirety). MS trials currently report efficacy as the relative difference in the proportion of patients assigned to control arms who progressed by trial's end and the proportion of patients assigned to active treatment arms who progressed by trial's end (CDP%Δ). The dynamic aspect of how patients achieve this outcome is ignored. As depicted in the Supplementary Figure 5A, the dynamic aspect of a DMT's therapeutic effect may inform its mechanism of action and predict the duration of therapeutic effect beyond the trial duration. We observed that, if a trial is of sufficient size, the proportion of patients assigned to placebo arms achieving A-CDP remains constant within trial duration. In other words, placebo arms tend to progress linearly, at least for duration of MS trials. Supplementary Figure 5A shows 3 hypothetical clinical trials of the same duration, with identical control arms achieving identical inhibition of annualized disability progression (i.e., same A-CDP%Δ). The difference among these trials is the dynamic aspect of the treatment effect: in the first trial (left panel), the efficacy remains constant throughout the trial. This suggests that treatment prevents accumulation of disability in the proportion of treated patients, at least during the trial. We observed this dynamic relationship between control and active treatment arms in trials that recruited early RRMS patients of young age and mild disability and treated them for up to 2 years. In the second trial (middle panel), the efficacy is maximal very early in the trial but decreases with trial duration as both arms tend to accumulate disability in parallel. This type of dynamic treatment effect is best exemplified by ORATORIO clinical trial of ocrelizumab in PPMS[1] and suggests that a drug only delays disability progression in some treated patients. Finally, the third theoretical trial (right panel) has delayed onset of the therapeutic effect, and the efficacy increases with trial duration. This dynamic effect suggests repair-promoting or neuroprotective mechanism of efficacy. We did not observe this dynamic effect among MS DMTs.

#### Supplementary Figure 5: Using dynamic analysis of treatment effect to develop more realistic efficacy outcome (i.e., A-CDP*TD%Δ) that assumes that MS DMTs on average only delay, rather than prevent disability progression

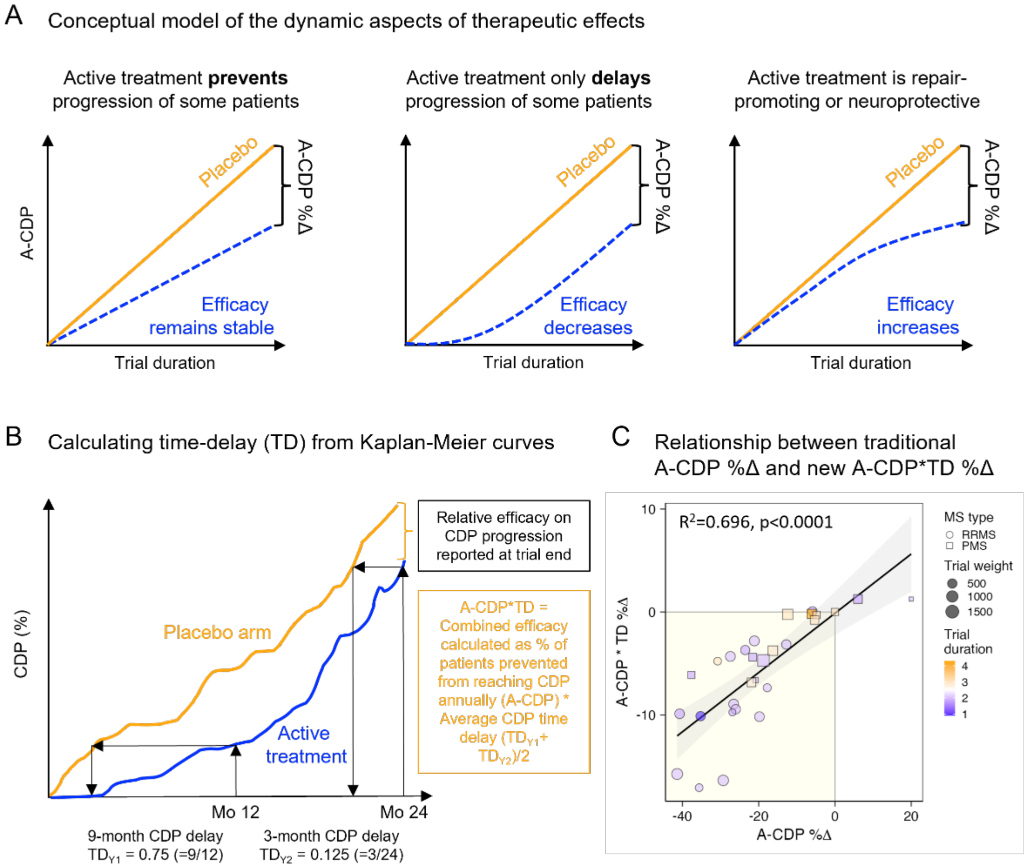

**Supplementary Figure 5**

**A.** Conceptual model depicting the importance of the dynamic aspect of the treatment effect on the hypothetical example of 3 clinical trials that have equivalent behavior of the control arm, same trial duration and reported identical traditional efficacy outcome (i.e., relative difference in the proportion of patients that progressed by trial' end; CDP%Δ). Because the trials have identical duration, the efficacy on annualized disability progression (A-CDP%Δ) is also identical. However, the different dynamic aspects of the therapeutic effect (i.e., stable, decreasing or increasing efficacy) provide important information about mechanism of treatment effect and potential efficacy of the drug beyond trial duration. **B.** Schematic depiction on how time-delay (TD) variables were generated from KM curves for each year of trial duration and how the average TD variable for years 1 and 2 of trial duration was used as modifier of efficacy to derive more realistic efficacy estimate (A-CDP*TD%Δ), that assumes that MS DMTs on average only delay, rather than prevent disability progression. **C.** Correlation between two efficacy outcomes: more traditional (and less realistic) A-CDP%D and TD-modified A-CDP*TD%D that assumes that MS DMTs on average only delay, rather than completely prevent disability progression. The $R^2$ and the p-value of the weighted Pearson correlation are depicted on the plot. RRMS trials are shown as circles, progressive MS trials as squares, with size proportional to trial weight. Trial duration is represented with a heatmap.

To formally measure dynamics of the therapeutic response in clinical trials that published K-M survival curves of sustained disability progression, we uploaded the copied survival curve image into Adobe Photoshop; then, we used an image analysis function and defined custom measurement scale (i.e., Image → Analysis → Set Measurement Scale → Custom) using the existing y axis scales, which allowed us to derive quantitative measurements of the proportion of subjects progressing at any time interval. Next, we created lines perpendicular to the x axis at each 1 year of the study, using Photoshop angular measurement to assure $90^o$ line angle to the x-axis. Where these yearly lines crossed active treatment and control arms of the KP curves, we created new lines parallel to x-axis. We applied the ruler tool with the "record measurement" command using the custom scale we defined above. This generated numerical values for proportions of patients progressing each year, which was inputted for some (older) trials that did not report these raw data (e.g., Cy1 trial).

We then analogously defined the custom measurement scale for the x-axis (trial duration) in months. Using this quantitative time axis, we asked by how much time (in months) the active treatment delays disability progression (Supplementary Figure 5B). To answer this question, we projected a proportion of patients that reached sustained disability progression on active treatment every year. E.g., in the A1 trial, 2.3% of alemtuzumab-treated patients reached sustained disability progression first year. We then asked when did 2.3% of control arm patients reach sustained disability progression and noted that it happened already at 3 months. So alemtuzumab therapy delayed disability progression by 9 months (75%; or time delay $[TD]_{y1} = 0.75$) in the first year of therapy. We generated analogous TD estimates for each subsequent year of the trial duration.

We focused on the first 2 years of Trial Duration because few trials were longer. Although initially we hoped to compare TD between first ($TD_{Y1}$) and second ($TD_{Y2}$) trial years, the trials that recruited early MS patients had too few progressing patients to make comparisons reliable. As $TD_{Y1}$ and $TD_{Y2}$ were correlated (i.e., weighted $r_{Pearson} = 0.745$, $R^2=0.555$, $p<0.0001$), we calculated mean $TD_{(Y1+Y2)/2}$ as a more reliable assessment of the DMTs' differences in the dynamic treatment effect.

The correlation between A-CDP%Δ and $TD_{(Y1+Y2)/2}$ from placebo-controlled trials of at least 2-year duration was significant but modest ($R^2 = 0.386$, $p=0.0016$), indicating that TD provides non-redundant information. Therefore, we used $TD_{(Y1+Y2)/2}$ as a modifier of reported efficacy to derive A-CDP*TD%Δ outcomes that assume that MS DMTs on average only delay rather than prevent disability progression. Expectedly, A-CDP%Δ and A-CDP*TD%Δ correlated (weighted $r_{Pearson} = 0.834$, $R^2=0.696$, $p<0.0001$; Supplementary Figure 5C) but A-CDP*TD%Δ showed much lower efficacies of MS DMTs. We believe that A-CDP*TD%Δ is a more realistic estimate of long-term, cumulative DMT efficacy.

##### Penalization function for trials that biased inclusion criteria against active comparator

11 out of 19 (56%) active-comparator trials that reported efficacy on CDP did not exclude patients who were previously treated with the active comparator. Because these trials simultaneously required evidence of disease activity in the inclusion criteria (i.e., requiring either relapse and/or CELs) they effectively enriched their studied population with active comparator non-responders. The proportion of recruited subjects previously treated with an active comparator ranged from 1.7 to 100%. We excluded from further analyses clinical trials that recruited 100% of active comparator non-responders (SENTINEL and CARE-MS II), as these trials studied de-facto different questions: whether switching non-responder to a new drug (CARE-MS; studied alemtuzumab) or adding a new drug to the non-responding drug (SENTINEL; studied natalizumab) improves efficacy. For both trials, the answer was affirmative: switching non-responders to a new drug is beneficial, as is adding a new drug (although this increases cost and side effects, and is not necessary). For the remaining 9 trials we devised a penalization function. A penalization function assumes that recruited active-comparator non-responders will continue to progress on an active comparator, effectively decreasing the predicted efficacy of the active comparator against placebo.

For example, in the D2 trial (DECIDE; studied daclizumab) 34.0% of patients randomized to the active comparator (i.e., Avonex) were previously treated with IFN-β preparations. If all patients were treatment naïve, in the recruited population IFN-β drugs are predicted to have -19.31% efficacy on A-CDP against placebo (i.e., Eq#6-predicted A-CDP%Δ = -20.95%, adjusted for weighted residual of IFN-β drugs = +1.64). However, 34.0% of recruited patients previously failed IFN-β drugs by fulfilling DECIDE trial inclusion criteria requiring minimum of 1 relapse/year. Penalization function assumed that 34.0% of patients will continue to progress on Avonex, which decreases predicted efficacy of Avonex against "in silico placebo" from -19.31% to -12.74%. With 8.6% of Avonex-treated patients reaching A-CDP in the DECIDE trial, the predicted -12.74% A-CDP%Δ of Avonex against placebo in D2 recruited population predicts A-CDP = 9.856% in the "D2 in-silico placebo arm". As A-CDP in daclizumab-treated patients in the D2 trial was 7.01%, the A-CDP%Δ of daclizumab against in-silico placebo is -28.9% (i.e., the difference between 7.01% and 9.856% progression).

#### Supplementary results

##### Study population

Among 61 randomized, blinded, and controlled Phase 2b or Phase 3 clinical trials, 42 (69%) enrolled only subjects with relapsing-remitting MS (RRMS), 10 enrolled only secondary-progressive MS (SPMS; 16%), 5 enrolled only primary-progressive MS (PPMS; 8%) and the remaining 4 enrolled both PPMS and SPMS (progressive MS [PMS]; 7%).

##### Comparative efficacy of MS DMTs, expanded results

Twenty-four clinical trials used an active comparator instead of placebo and all studied RRMS. Most (17; 71%) compared efficacy of the newer drug to one of the interferon-β products, five (21%) to teriflunomide one (4%) to dimethyl fumarate and one (4%) to glatiramer acetate. Only 19 (80%) active comparator trials reported efficacy on CDP.

Alemtuzumab trials used only rater-blinded design. Patients knew their trial tested alemtuzumab's superiority and they knew their assigned treatment. As patients' motivation influences disability measurements (especially for EDSS<4) this design likely over-estimates alemtuzumab's efficacy.

We mathematically predicted "in-silico placebo arms" for these active comparator trials as described in Methods and Supplementary Methods, in order to use these trials for comparative efficacy of MS drugs (Figure 2G), based on residuals from Eq#6-predicted and measured efficacies against placebo. We saw that these residuals were not uniformly distributed but were highest for smallest trials (and among trials with weight<700 were 3 outliers) and as trial size increased, the residuals decreased (Supplementary Figure 8). We conclude that imprecision (highest in the small trials) explains some residual variance.

#### Supplementary Figure 6: Smaller trials have larger residuals from observed versus Eq#6-predicted efficacy

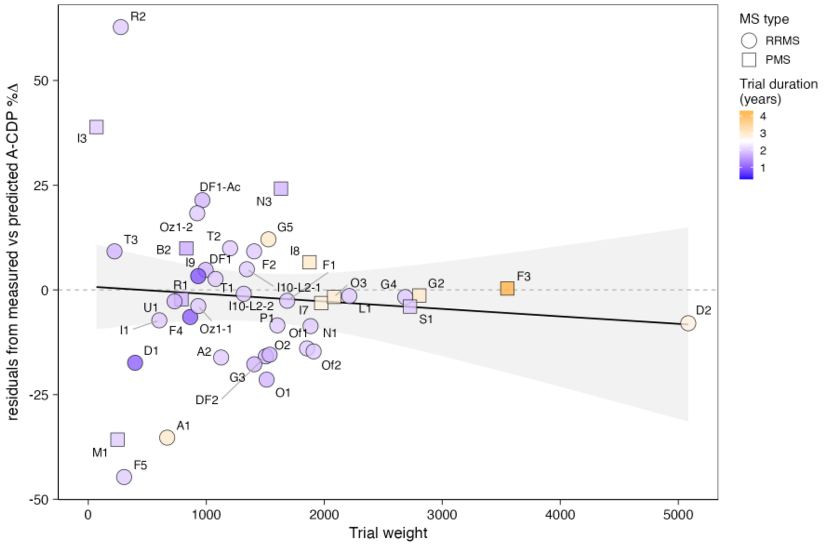

**Supplementary Figure 6:**

Funnel-like distribution of the residuals from observed-versus Eq#6-predicted efficacy rates dependent on trial weight ($n * \sqrt{TrialDuration}$) is consistent with imprecision of smaller trials. Based on this distribution we consider trials with weight <700 to be unreliable in their efficacy estimates. RRMS trials are depicted as circles and PMS trials as squares. Trial duration is depicted as color with shorted duration in blue and longer duration in orange. The trial name corresponds to Index column in the Supplementary Master Worksheet (SMW).

#### Risk of MS treatments, expanded results

The retrospective study of Kaiser Permanente members spanning 12 years (2008-2020) included 6,626 MS patients with 11,929 treatment episodes and compared them to 33,550 age, sex, race, and ethnicity matched population controls[2]. The Swedish nationwide register-based study spanning 6 years (2011-2017) included 6,421 MS patients with 8,600 treatment episodes and 42,645 equally matched population controls[3].

The results of these studies overlapped. The USA cohort demonstrated an increase in adjusted risk ratio (aRR) of all outpatient infections even in untreated MS patients (aRR=1.39). This ratio increased in MS patients treated with IFNβ/GA (aRR=1.6). Both cohorts saw additional infection increases in patients that were treated with newer DMTs (i.e., rituximab, natalizumab, fingolimod; aRR between 1.73-1.99).

Even more concerning was almost 3 times higher risk of serious infections in untreated MS (aRR = 2.97). While the interferon-β/glatiramer acetate treatments did not increase the risk of serious infections further (aRR – 2.31), the newer drugs did in both cohorts, reaching up to aRR 4.34 for natalizumab.

Investigating predictors of serious infections in the MS cohort identified the significant effects of age, comorbidities, and advanced disability. Every decade of age increased the hazard ratio (HR) of serious infections in MS patients by 1.36. Charlson Comorbidity Index[4] (CII) of 1 increased this HR by 1.46. CII 1 is assigned to patients with following single comorbidities: myocardial infarction, congestive heart failure, cerebrovascular disease, dementia, chronic pulmonary disease, ulcer disease, mild liver disease and diabetes. Patients with any combination of these comorbidities, a higher grade of liver disease or diabetes, or any other serious disease (such as cancers) have CII of ≥ 2, causing their severe infection HR to jump to 3.87. Most concerning, the HR of serious infections for patients with EDSS > 6 (i.e., non-ambulatory or requiring bilateral support for ambulation) is 5.29, irrespective of comorbidities or age.

The effect of MS DMTs on cancer morbidity/mortality is more difficult to estimate due to inadequate/conflicting data. All identified studies focused on de-novo cancer risk and included relatively short drug exposures (as short as 6 months); consequently, there is no data allowing estimation of long-term/cumulative risk of cancer in DMT-treated individuals. Neither is there data for predicting the effect of DMTs on cancer recurrence or secondary cancers.

With these limitations, we summarize published literature, focusing on results congruent between studies. Although sufficiently large population-level results are missing, the published studies generally find that cancer incidence in untreated MS is likely comparable to general population. The French nation-wide registry study identified 95,474 MS patients captured as unique incidence cases without prior history of cancer between 1/1/2008-12/31/2014 and matched these 1:1 with population controls[5]. MS patients had 1.36 times increased incidence of cancer (HR 95% CI = 1.29-1.43), across all age categories and genders. Lacking available data, the study did not match for smoking or obesity, which are associated higher incidence of both MS and cancer (lung with smoking, colorectal cancer with obesity). Follow-up study[6] identified a subpopulation of 28,720 cancer-naïve, newly treated MS patients and linked increased risk of cancer to DMT exposures: the study found 181 out of 19,146 individuals developing cancer on interferon-β and glatiramer acetate with identical odds ratio (OR) as a matched control population. Significantly fewer patients were treated with remaining drugs (i.e., dimethyl fumarate, teriflunomide, natalizumab, fingolimod and non-MS specific immunosuppressive drugs azathioprine, mycophenolate, methotrexate) and these drugs had an OR for cancer incidence of 1.36.

The observational, cross-sectional pharmacovigilance disproportionality analysis using World Health Organization database: VigiBase® found 5,955 cancer cases from 240,993 reports of MS DMTs.[7] After confounder adjustment, the study found a disproportional increase in cancer reports linked to natalizumab (OR 1.74), interferon-β (OR 1.39), dimethyl fumarate (OR 1.35) and fingolimod (OR 1.15). It did not find significant associations with glatiramer, teriflunomide, alemtuzumab and ocrelizumab, but due to the short time these drugs (other than glatiramer acetate) are on the international markets, there were too few reports in the VigiBase® to conclude that these drugs do not increase cancer incidence.

Swedish nationwide registries found borderline increased cancer risk with fingolimod, but not with rituximab and natalizumab[8]. The Danish MS registry did not find increased cancer risk with MS DMTs[9].

Unfortunately, these negative studies used "intention-to-treat" analysis, which means that they analyzed patients even if they switched therapy.

Despite these somewhat conflicting results, based on the MOA of MS DMTs and cancer immunosurveilance knowledge, it is highly probable that at least some DMTs increase cancer incidence with long-term exposure. Clearly longer population-based studies are required to better estimate cancer risks of MS DMTs.

#### Supplementary discussion

Some of the stated weaknesses of presented results, such as the lack of patient-level data forcing us to model only population averages, are also strengths of this comprehensive meta-analysis of DMT efficacy. For example, we observed that only 14.71% of MS patients progressed yearly in placebo-controlled trials yearly thanks to poor EDSS sensitivity. This progression rate over-estimates progression in general MS population because patients were pre-selected for high lesional activity (for relapse-onset MS) or high progression rates (for most PMS trials), in comparison with natural history cohorts, where patients progress approximately by 1 EDSS point per decade[10]. Exactly which at-risk patients progress during trial duration is somewhat stochastic; this makes analysis of patient-level data challenging. Instead, clinical trial averages transform outcomes and most predictors into normally distributed continuous variables measured with high accuracy. Nevertheless, access to patient level data would allow us to group patients into decade-delimited age categories, which would mitigate the problem of stochastic EDSS progression and likely yield stronger models, especially for comparative efficacies between MS DMTs.

Applying results derived from group averages to individuals may not be intuitive, but the proportion of patients reaching A-CDP on EDSS is equivalent to the mean of (EDSS-based) yearly progression slopes. E.g., A-CDP of 0.2 means that 20% of patients progressed on EDSS and 80% did not progress. This means that 80% of patients will have yearly EDSS-based progression slopes of 0 and 20% will have, on average, a slope of 1 (older patients may have slope of 0.5 depending on the employed definition of progression, while others may have a slope greater than 1 reflecting earlier or more severe progression). This is equal to the mean EDSS-based yearly progression slope of 0.2. Thus, even though the presented models reflect behavior of and "average" MS patient, they provide patient-level predictions.

Our study does not determine if a high efficacy DMT given at MS onset eliminates PILA development. Whether early aggressive treatments may be safely de-escalated to lower efficacy drugs (that don't increase infectious mortality) likely depends on the mechanism(s) behind the MS lesional activity decline observed in the placebo arms. If this decline is due to inflammation-driven establishment of tertiary lymphoid follicles (compartmentalized inflammation), lesional activity will likely reappear after cessation of treatments that block immune cells trafficking to CNS tissue such as natalizumab or fingolimod. On the other hand, if lesional activity depends on subpopulations of immune cells (e.g., B cells, perhaps EBV infected), then this activity may not reappear after a long treatment with the appropriate high-efficacy DMT (i.e., B cell depleting treatments, including alemtuzumab). These critical questions must be tested in new clinical trials or observational studies, some of which are currently ongoing under the sponsorship of PCORI.

Our results also question the ethics of using a weak active comparator in future clinical trials of RRMS patients: the presented data clearly predicts that "lost efficacy" cannot be recuperated by subsequently switching to a more efficacious drug and that DMT efficacy is highest when patients initiate high efficacy DMTs close to MS onset. Delaying such a high efficacy DMT, even for several years of trial duration, is not in the patients' best interest.

Comparing efficacy between MS DMTs (Figure 4G) showed some surprising observations. For example, we found that less immunogenic B cell-depleting monoclonal antibodies (ocrelizumab and ofatumumab) have higher efficacy compared to chimeric (and therefore more immunogenic) antibodies ublituximab and rituximab. Notably, the real-world comparison of ocrelizumab and rituximab supports our conclusion of ocrelizumab's superiority[11]. Likewise, for drugs that target S1P receptors (S1PR), we observed the following efficacy hierarchy: ponesimod (binds to S1PR1 only) > Siponimod (binds S1PR1 and S1PR5) > fingolimod (nonspecific binding to multiple S1P receptors). (We already noted that ozanimod, which targets both S1PR1 and S1PR5 has discrepancy in comparative efficacy between doses, although the lower dose had predicted efficacy completely overlapping with siponimod, another S1PR1&5 targeting drug). This hierarchy suggests that binding S1P receptors other than S1PR1 limits efficacy. It is intriguing to note that S1PR1 (and S1PR4), but not S1PR5, -2 or -3 are expressed in B cells. The broad expression levels of these receptors in non-immune cells indicates that S1PR-targeting drugs may have many unexpected effects, including on mesenchymal cells involved in processes such as fibrosis, especially if they are non-specific, such as fingolimod.

Comparative efficacy analyses also reveal that most small trials are efficacy outliers; they provide unrealistically high (M1; mitoxantrone trial, A1 alemtuzumab trial) or unrealistically low (I3 interferon-β trial and R2 rituximab trial) efficacy estimates, likely because too few patients reach a sustained disability progression on EDSS. Furthermore, the evidence that DMTs that share mechanisms of action (i.e., ocrelizumab and ofatumumab, ponesimod and siponimod, rituximab and ublituximab) also cluster together in the comparative efficacy chart strengthens our confidence in the hierarchy of comparative efficacies. That being said, the comparative efficacy data are not amenable to formal statistical analysis, and we note that comparative clinical trials will never be performed due to impractically low power of reasonably sized trials. Comparative differences between MS DMTs on MS disability progression are too small to be identified even in multicentric real-world observational studies such as MSbase[11-16]. Therefore, comparative efficacy should not be the only determining factor when selecting a DMT; the side-effect profile must be also considered. In this regard, we highlight critical need to assess infectious risk of newer DMTs and, most importantly, long-term risks of all MS DMTs on incidence and severity of primary and secondary cancers.
